# Genome-wide association analysis reveals insights into the molecular etiology underlying dilated cardiomyopathy

**DOI:** 10.1101/2023.09.28.23295408

**Authors:** Sean L Zheng, Albert Henry, Douglas Cannie, Michael Lee, David Miller, Kathryn A McGurk, Isabelle Bond, Xiao Xu, Hanane Issa, Catherine Francis, Antonio De Marvao, Pantazis I Theotokis, Rachel J Buchan, Doug Speed, Erik Abner, Lance Adams, Krishna G Aragam, Johan Ärnlöv, Anna Axelsson Raja, Joshua D Backman, John Baksi, Paul JR Barton, Kiran J Biddinger, Eric Boersma, Jeffrey Brandimarto, Søren Brunak, Henning Brundgaard, David J Carey, Philippe Charron, James P Cook, Stuart A Cook, Spiros Denaxas, Jean-François Deleuze, Alexander S Doney, Perry Elliott, Christian Erikstrup, Tõnu Esko, Eric H Farber-Eger, Chris Finan, Sophie Garnier, Jonas Ghouse, Vilmantas Giedraitis, Daniel F Guðbjartsson, Christopher M Haggerty, Brian P Halliday, Anna Helgadottir, Harry Hemingway, Hans Hillege, Isabella Kardys, Lars Lind, Cecilia M Lindgren, Brandon D Lowery, Charlotte Manisty, Kenneth B Margulies, James C Moon, Ify R Mordi, Michael P Morley, Andrew D Morris, Andrew P Morris, Lori Morton, Mahdad Noursadeghi, Sisse R Ostrowski, Anjali T Owens, Colin NA Palmer, Antonis Pantazis, Ole BV Pedersen, Sanjay K Prasad, Akshay Shekhar, Diane T Smelser, Sundarajan Srinivasan, Kari Stefansson, Garðar Sveinbjörnsson, Petros Syrris, Mari-Liis Tammesoo, Upasana Tayal, Maris Teder-Laving, Guðmundur Thorgeirsson, Unnur Thorsteinsdottir, Vinicius Tragante, David-Alexandre Trégouët, Thomas A Treibel, Henrik Ullum, Ana M Valdes, Jessica van Setten, Marion van Vugt, Abirami Veluchamy, W.M.Monique Verschuuren, Eric Villard, Yifan Yang, COVIDsortium, DBDS Genomic Consortium, Genomics England Research Consortium, HERMES Consortium, Folkert W Asselbergs, Thomas P Cappola, Marie-Pierre Dube, Michael E Dunn, Patrick T Ellinor, Aroon D Hingorani, Chim C Lang, Nilesh J Samani, Svati H Shah, J Gustav Smith, Ramachandran S Vasan, Declan P O’Regan, Hilma Holm, Michela Noseda, Quinn Wells, James S Ware, R Thomas Lumbers

**Author notes:** Contributed equally (Sean L Zheng, Albert Henry). Full list in Supplementary Materials (HERMES Consortium). Contributed equally (James S Ware, R Thomas Lumbers).

## Abstract

Dilated cardiomyopathy (DCM) is a clinical disorder characterised by reduced contractility of the heart muscle that is not explained by coronary artery disease or abnormal haemodynamic loading. Although Mendelian disease is well described, clinical testing yields a genetic cause in a minority of patients. The role of complex inheritance is emerging, however the common genetic architecture is relatively unexplored. To improve our understanding of the genetic basis of DCM, we perform a genome-wide association study (GWAS) meta-analysis comprising 14,255 DCM cases and 1,199,156 controls, and a multi-trait GWAS incorporating correlated cardiac magnetic resonance imaging traits of 36,203 participants. We identify 80 genetic susceptibility loci and prioritize 61 putative effector genes for DCM by synthesizing evidence from 8 gene prioritization strategies. Rare variant association testing identifies genes associated with DCM, including *MAP3K7, NEDD4L*, and *SSPN*. Through integration with single-nuclei transcriptomics from 52 end-stage DCM patients and 18 controls, we identify cellular states, biological pathways, and intercellular communications driving DCM pathogenesis. Finally, we demonstrate that a polygenic score predicts DCM in the general population and modulates the penetrance of rare pathogenic and likely pathogenic variants in DCM-causing genes. Our findings may inform the design of novel clinical genetic testing strategies incorporating polygenic background and the genes and pathways identified may inform the development of targeted therapeutics.

---

Dilated cardiomyopathy (DCM) describes a spectrum of heart muscle diseases that are characterized by ventricular dilatation and/or impaired myocardial contractility in the absence of coronary artery disease or abnormal loading conditions^1,2^. DCM affects ∼1/250 individuals and is one of the primary aetiologies of heart failure and the leading cause of cardiac transplantation^3^. Pathogenic mutations in relevant genes can cause DCM via monogenic disease mechanisms, however, recent evidence suggests individuals’ polygenic background can modify the risk of developing DCM^4^. Characterization of the genetic architecture underlying DCM provides opportunities for improved clinical genetic testing and the discovery of pathways and genes to inform therapeutic development and improve patient outcomes.

We performed a meta-analysis of case-control DCM GWAS comprising 14,255 cases and 1,199,156 controls from 16 studies participating in the Heart Failure Molecular Epidemiology for Therapeutic Targets (HERMES) Consortium^5^ (GWAS_DCM_; **Figure 1, Tables S1-2, Supplementary Methods**). Of the 16 studies, 6 ascertained cases using strict criteria which require cardiac imaging, specific ICD diagnostic codes, and/or a clinical diagnosis of DCM (DCM_Strict_: 6,001 cases and 449,382 controls), while 10 ascertained cases based on left ventricular systolic dysfunction (LVSD) in the absence of secondary causes (DCM_Broad_: 9,298 cases and 1,157,145 controls). Of 9,656,392 common variants (minor allele frequency [MAF]>0.01) included in the meta-analysis, we identified 63 conditionally independent (sentinel) variants at 62 genomic loci passing a false discovery rate (FDR) <1%, including 27 sentinel variants at 26 loci passing genome-wide significance (*P*<5x10^−8^) (**Figure 2, Figure S1, Table S3**).

**Figure 1:**
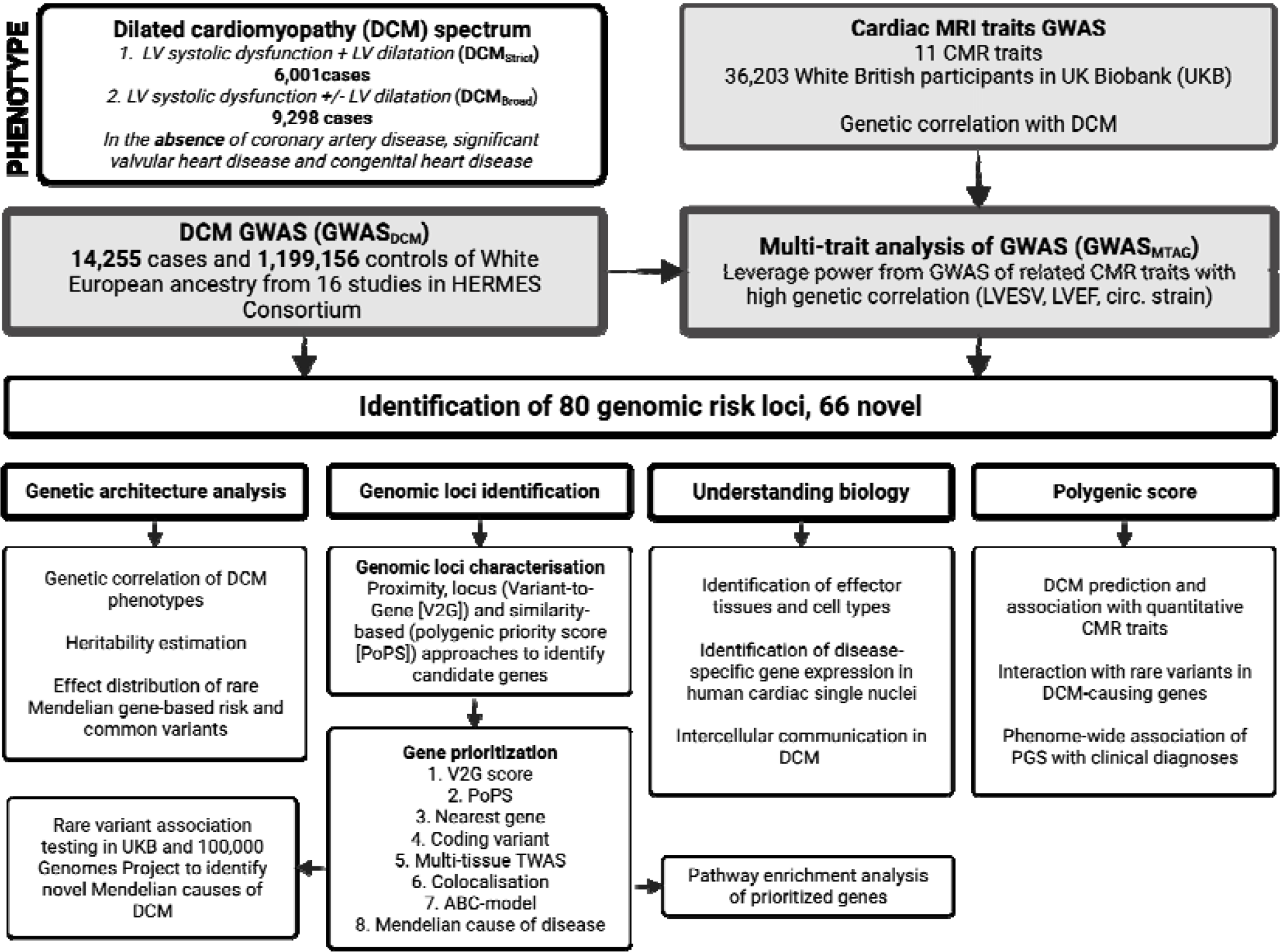
Study overview of European Ancestry GWAS of dilated cardiomyopathy spectrum (GWAS_DCM_) performed in 14,256 cases and 1,185,671 controls from 16 studies. Cases were defined as having LV systolic dysfunction with or without LV dilatation (DCM_Broad_), or LV systolic dysfunction and LV dilatation (DCM_Strict_) in the absence of coronary artery disease, significant valvular heart disease or congenital heart disease. Multi-trait analysis of GWAS (MTAG) was performed combining GWAS_DCM_ with GWAS of genetically-correlated quantitative cardiac CMR traits (GWAS_MTAG_). Genomic risk loci were identified and systematically annotated to prioritize candidate genes at each locus.

**Figure 2:**
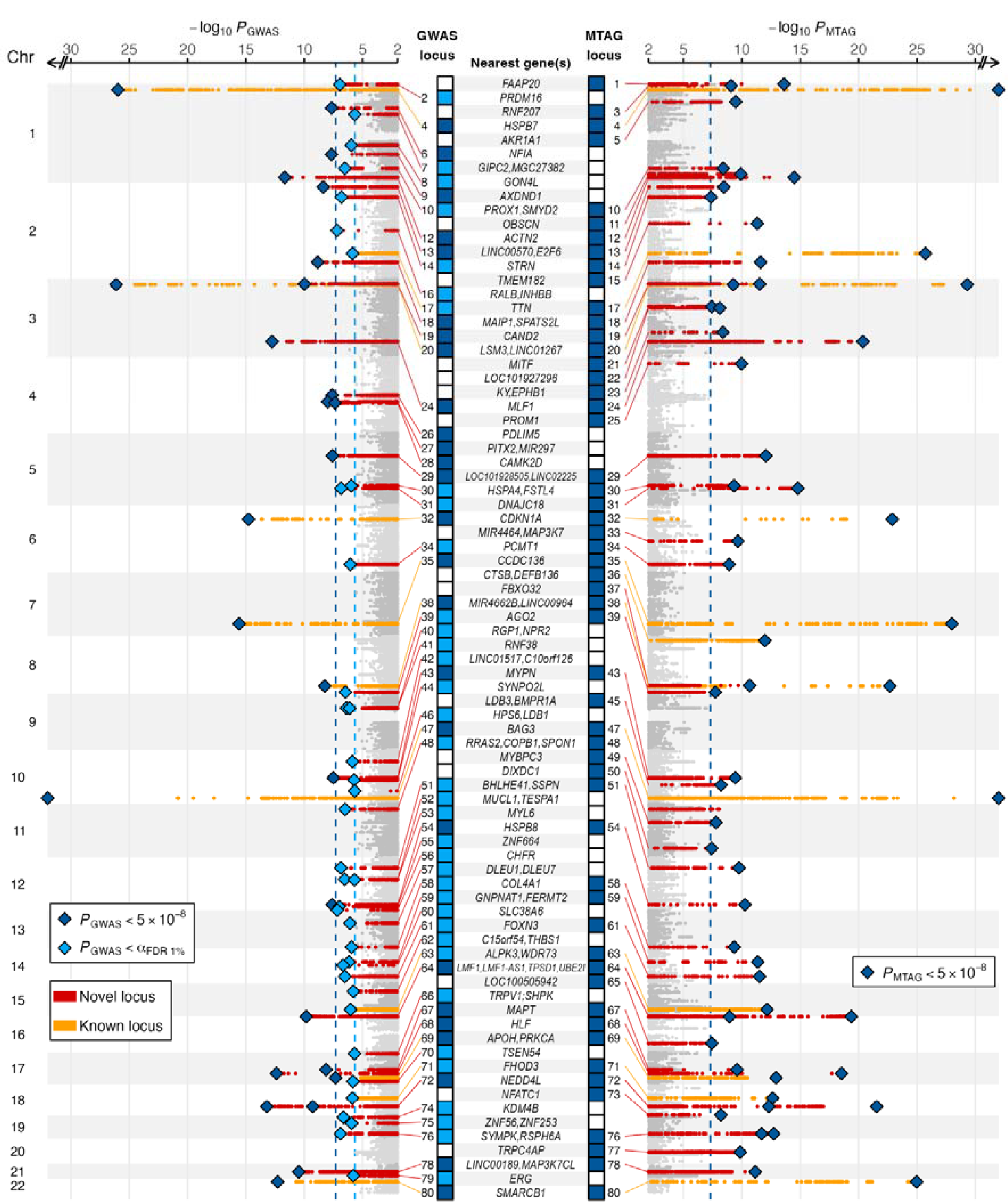
Manhattan plot of primary GWAS and MTAG identifying novel (red) and previously reported^8,37^ (orange) genomic loci associated with DCM. Loci reaching genome-wide (P<5x10-8, blue) in GWAS and MTAG, and FDR (alphaFDR0.01, light blue) in GWAS are highlighted. Loci are annotated with the nearest gene(s) of all conditionally independent variants within the locus and ordered in ascending genomic location.

To assess differences in genetic association signals that may arise from ascertainment of DCM cases, we performed a GWAS meta-analysis of the 6 studies that used strict case definitions (GWAS_DCM-Strict_) (**Figures S1 and S2**). In this subset, all lead variants within 62 loci identified in GWAS_DCM_ showed a concordant direction of effect, including 10 loci with one or more lead variants passing *P*<5x10^−8^ (**Table S3**). Most sentinel variants showed a larger effect in GWAS_DCM-Strict_ (**Figure S3** and **S4**). Using linkage-disequilibrium adjusted kinships (LDAK) with summary statistics from the GWAS meta-analysis^6^, we estimated the heritability explained by common SNPs (*h*^*2*^_*SNP*_) on the liability scale as 20% (2.1% SD) for GWAS_DCM-Strict_ and 11% (1% SD) for GWAS_DCM_. Using linkage disequilibrium score regression (LDSC), we estimated the heritability explained by common genetic variants (h_g_^2^) on the liability scale as 23% for GWAS_DCM-Strict_ and for 11% for GWAS_DCM_. We found a near complete genetic correlation between GWAS_DCM_ and GWAS_DCM-Strict_ (*r*_g_>0.99).

To explore shared genetic etiology with quantitative left ventricular (LV) traits, we estimated the pairwise genetic correlation (*r*_g_) of DCM with ten cardiac magnetic resonance imaging-derived traits from 36,203 participants in the UK Biobank (UKB) using bivariate LD-score regression^7,8^; LV end-systolic volume (LVESV) (*r*_g_ 0.73), circumferential strain (*r*_g_ 0.71) and ejection fraction (LVEF) (*r*_g_ -0.70) were highly correlated with DCM (**Table S4**). Based on these observations we included these traits in a DCM-anchored multi-trait analysis of GWAS (GWAS_MTAG_), enabling joint analysis to increase genome-wide statistical power^9^. Through GWAS_MTAG_, 58 sentinel variants at 54 loci were identified at *P*<5x10^−8^, including an additional 18 loci that were not identified in GWAS_DCM_ at FDR <1%. Overall across both GWAS_DCM_ and GWAS_MTAG_, there were a total of 80 genomic risk loci, of which 66 are novel associations with DCM (previously identified loci reported in **Supplementary Methods**). Regional association plots for all 80 risk loci are available in the Supplementary Material. Among loci from GWAS_DCM_, 25 FDR-significant loci were not significant in GWAS_MTAG_, although all uniquely significant loci (GWAS_DCM_ and GWAS_MTAG_) shared directional concordance (**Figures S3**-**S5**).

Using functionally-informed fine-mapping, we identified 100 credible sets of likely causal variants at 63 of 80 loci. The credible sets consist of 1,392 variants (60.6% intronic, 25.4% intergenic and 4.8% exonic), including 83 variants with a posterior inclusion probability (PIP) >0.5 identified at 43 loci (**Figure S6, Table S5**). Several fine-mapped coding variants were identified within known DCM genes (*FLNC, BAG3*, and *TTN*) and genes with plausible modifying effects on cardiac function (*NEXN* and *MYBPC3*), including deleterious missense variants (CADD Phred score>15) in *TTN, BAG3*, and *MYBPC3*.

Next, we prioritized effector genes for DCM by evaluating functional evidence of 1,970 protein-coding genes situated within or overlapping identified genomic risk loci (**Figure 3A, Table S6**). First, using a combination of nearest gene, locus-based (variant-to-gene [V2G]), and similarity-based (polygenic priority score [PoPS]) methods we identified 380 candidate genes for further prioritization (median 5 per locus [IQR 4 to 6]). Second, using evidence from 5 additional methods (coding variant, co-localization with expression quantitative trait loci [eQTL], transcriptome-wide association study [TWAS], activity-by-contact [ABC]-model, and established Mendelian cardiomyopathy or muscle-disease-causing genes) we prioritized a single high-confidence gene at 61 of 80 loci (**Figure 3B, Figure S7, Table S7**). The highest prioritization scores were for *MYPN* (prioritized by 7 out of the maximum of 8 predictors), followed by *HSPB8* and *ALPK3* (6/8), and *ACTN2, SPATS2L* and *BAG3* (5/8). Highlighting the robustness of this framework, all ClinGen cardiomyopathy-causing genes with definitive evidence of being Mendelian causes of cardiomyopathy except *LMNA*, together with several genes with limited evidence for cardiomyopathy (*LDB3, MYPN, PRDM16, OBSCN*), were prioritized at their respective loci. Genes that caused Mendelian forms of HCM (*MYBPC3, ALPK3, FHOD3*) were also identified at genomic risk loci for DCM; a finding consistent with prior evidence that these disorders represent opposing extremes of a continuum of ventricular structure and systolic function^8,10^.

**Figure 3:**
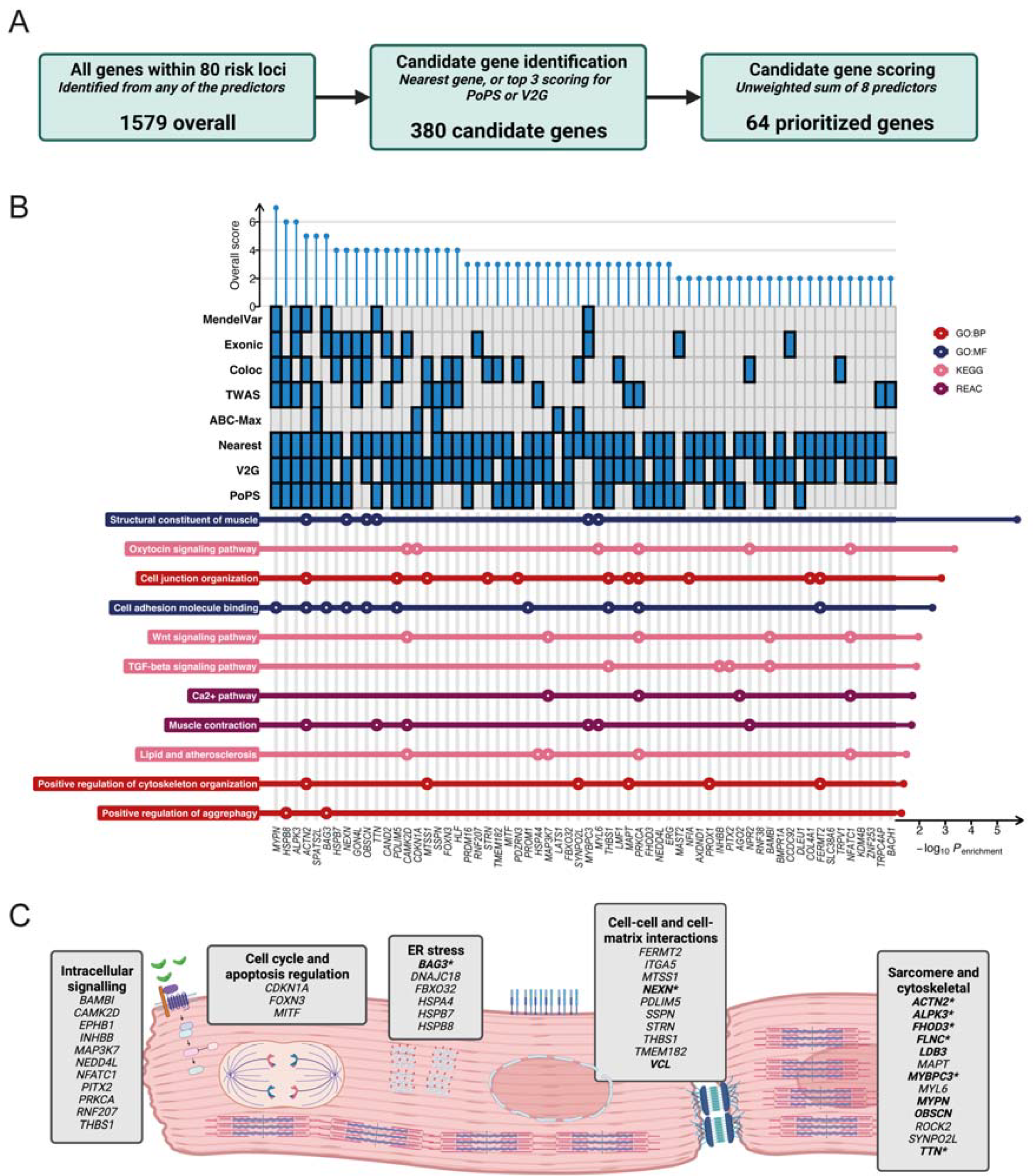
Locus annotation and candidate gene scoring prioritize genes at risk loci and important biological pathways and processes in DCM pathogenesis. (A and B) Among all genes located within genomic risk loci (1,970 genes), candidate genes were selected based on proximity and being one of the top 3 genes predicted using V2G or PoPS (380 candidate genes). 61 genes were prioritized at 61 loci after scoring highest among the 8 predictors. (B) Pathway enrichment analysis of prioritized genes highlighted pathways related to muscle structural constituents. (C) Schematic overview of pathways and processes highlighted in DCM pathogenesis. Manually curated from pathway enrichment analysis and published literature.

Pathway analysis of prioritized genes identified enrichment of 72 biological pathways, including sarcomeric and cytoskeletal function, cellular adhesion and junction organization, aggrephagy, Wnt and TGF-β signaling (**Figure 3B** and **3C, Table S8**). Among the novel genes identified in these analyses were additional genes with contractile and cytoskeletal functions, including *MAPT*^*11*^ and *MYL6*^*12*^. The important role of cell-to-cell adhesion and cell-to-matrix interaction in DCM pathogenesis was highlighted by many effector genes acting at these interfaces. *STRN* encodes the desmosomal protein striatin, the canine ortholog of which has been implicated in dilated and arrhythmic cardiomyopathy^13^. *SSPN* encodes sarcospan, a key component of the dystrophin glycoprotein complex (DGC) that has been linked to severe skeletal and cardiac muscle disorders. Other effector genes acting at the cell membrane identified include *MTSS1*^*14*^, *PDLIM5*^15,16^, *THBS1*, and *TMEM18*^17^.

Cell signaling components were prominent among the prioritized genes, including members of the TGF-β (*BAMBI, INHBB, PITX2*, and *THBS1*) and Wnt signaling pathways (*CAMK2D, MAP3K7, NEDD4L, NFATC1, PRKCA*, and *RNF207*). *INHBB* encodes a secreted factor and *THBS1* a transmembrane glycoprotein, both of which activate the TGF-β receptor, while *BAMBI* encodes a TGF-β-like pseudoreceptor that acts as a negative regulator of TGF-β signaling^18^. TGF-β signaling has been shown to be important in the development of fibrosis in cardiomyopathy models^19^. Finally, expanding upon the established role of *BAG3* and the unfolded protein response and endoplasmic reticular stress on DCM pathogenesis, several genes encoding heat-shock proteins (*HSPA4, HSPB7*, and *HSPB8*) were identified. In addition, *FBXO32* encodes a muscle-specific ubiquitin ligase involved in protein degradation that has been postulated to be a rare cause of DCM^20^.

For genomic loci where a single high-confidence genes could not be identified, we manually curated the locus by integrating information from enriched biological pathways. The identified candidate genes map to cytoskeleton function (*ROCK2*^*21*^ at locus 13), cell adhesion (*ITGA5* at locus 52), MAPK signaling (*EPHB1* at locus 23), and the unfolded protein response (*DNAJC18* at locus 31, *CRYAB* at locus 50). Other notable genes included the taurine transporter *SLC6A6* (locus 20) with existing evidence of taurine-deficiency causing feline DCM^22^, the cardiac-expressed K+ channel *KCNIP2* that has been implicated in Brugada syndrome and conduction abnormalities^23^, *RRAS2* where gain of function variants are a cause of Noonan syndrome and accompanying hypertrophic cardiomyopathy^24,25^, and several genes implicated in myopathy (*CHCHD10* at locus 80, and *DMPK* at locus 76.

Within the identified DCM loci were 7 Mendelian cardiomyopathy genes catalogued in ClinGen with definitive evidence (DCM: *TTN, FLNC, LMNA, BAG3*, HCM: *MYBPC3, ALPK3, FHOD3*) and 7 moderate or limited-evidence genes (DCM: *PRDM16, LDB3*; DCM or HCM: *OBSCN, VCL, NEXN, MYPN*; intrinsic cardiomyopathy: *ACTN2*). Emphasizing the role of gene dosage as a likely mechanism of action at GWAS genes^26^ and continuum of disease risk, 4 of the 6 DCM genes established to act through mechanisms that include reduced gene product^27^ were identified through GWAS (*TTN, FLNC, LMNA*, and *BAG3*), with a 10-fold enrichment of Mendelian cardiomyopathy genes within GWAS loci (OR=10.0, *P=*1.1x10^−6^).

We next performed rare variant (MAF<0.001) burden testing of (1) all ClinGen definitive and moderate evidence DCM-causing genes^28^ (predicted truncating variants [PTV] or missense variants) to characterize the overall genetic architecture of DCM, and (2) candidate genes identified at GWAS locus through functional genomics analysis (PTVs only) to identify novel Mendelian causes of DCM and cardiomyopathy. In 453,455 participants with whole exome sequencing (WES) in UKB, a population-based cohort recruiting middle-aged and older individuals with no disease focus, the combined risk effect of rare variants in ClinGen definitive or moderate evidence DCM genes were orders of magnitude higher than GWAS sentinel variants mapping to the same genes (**Figure 4A, Table S9**).

**Figure 4:**
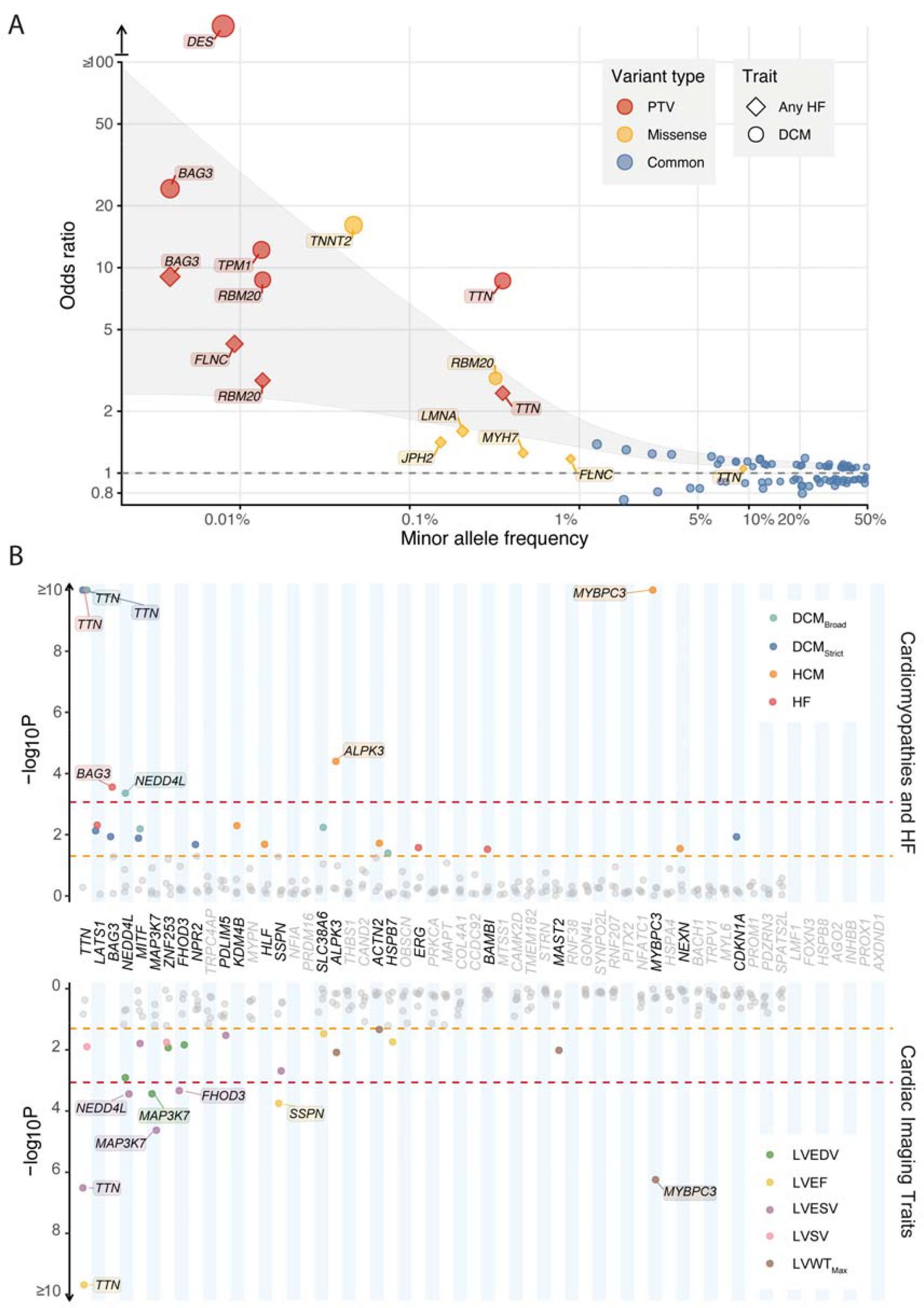
Rare variant analysis highlights the genomic architecture of dilated cardiomyopathy and identifies novel disease- and trait-associated genes. (A) Burden testing was performed for rare variants (MAF<0.001) in genes with moderate or definitive evidence of causing DCM^28^, collapsed into 2 variant classes: predicted truncating (PTV, red), and missense (yellow) variants. Individual sentinel common variants (MAF>0.01) in DCM loci are highlighted in blue. Variant frequency represents MAF for individual sentinel variants, and the cumulative population frequency of rare variants in burden tested genes. Outcome for burden testing were DCM and heart failure, with all gene masks reaching nominal significance (P<0.05) presented. The grey highlighted region indicates smoothened regression lines of the upper and lower bounds for each effect estimate. (B) Burden analysis of rare PTVs (MAF<0.001) in 58 prioritized protein-coding genes in UKB (453,455 participants with WES, and 36,104 with CMR) highlights known Mendelian disease-causing genes (*TTN, BAG3, FHOD3, ALPK3* and *MYBPC3*) and 3 novel genes (*NEDD4L, MAP3K7*, and *SSPN*). Red line indicates statistical significant (P<8.6x10^−4^ [0.05/58 genes]) and orange line indicates nominal significance (P<0.05). Genes are ordered by mean P-value across all tested traits, from lowest to highest, with genes reaching nominal significance (P<0.05) for at least one trait highlighted in bold. Detailed results available in **Tables S11-13**.

To identify genes with novel Mendelian causes of DCM and cardiomyopathy more generally, we evaluated the effect of deleterious rare variants in the 61 prioritized genes with binary (cardiomyopathy and heart failure phenotypes) and quantitative CMR traits. Analysis was performed using whole genome data in 78,142 individuals participants of Genomics England (GeL), a rare disease and cancer cohort that recruited probands and their relatives from clinical centers, and with WES in the 453,455 participants (including a subset of 36,104 with CMR) in the UKB. PTVs in three genes which currently lack sufficient evidence to be definitive causes of Mendelian disease were nominally found to be associated with DCM (*MYPN*: OR 15.0, P 0.03; *PRDM16*: OR 40.3, P 0.008) in GeL, and with HCM (*NEXN*: OR 24.1, P 0.01) in UKB.

Given our finding of widespread allelic series for DCM genes, we explored genomic loci that did not harbor genes with assertion for autosomal DCM. Rare PTVs in 3 prioritized genes that are not established causes of cardiomyopathy were found to be associated with binary diseases outcomes (*MAP3K7* and *NEDD4L* with DCM) in at least one cohort (**Figure 4B, Table S10** and **S11**), and with quantitative traits (*NEDD4L, MAP3K7*, and *SSPN*) in UKB (**Figure 4B, Table S12**). PTVs in *MAP3K7* were associated with DCM in GeL (OR 24.2, P_adj_ 0.02), and increased LV volumes (LV end-diastolic volume [LVEDV] +54ml, P_adj_ 0.01, LVESV +38ml, P_adj_ 4.4x10^−4^) in UKB. The importance of *MAP3K7* in DCM pathogenesis is highlighted by prioritization of additional pathway genes, including *RNF207*^*29*^, a regulator of the *MAP3K7* activator *TAB1*. PTVs in the membrane receptor regulator *NEDD4L* were associated with DCM in UKB (OR 10.4, P_adj_ 0.01), and HF in GeL (HF OR 13.0, P_unadj_ 0.01), and quantitative traits (PTV: LVEDV +29.7, P_adj_ 0.02; LVESV +19.8, P_adj_ 0.005). PTVs in *SSPN* were associated with significant changes in quantitative LV traits (LVEF -5.9%, P_adj_ 0.004 and LVESV +13.0ml, P_adj_ 0.02). Within a local DCM cohort, 3 of 337 cases (0.9%) carried PTV in *SSPN* compared with 80 in 352,564 (0.02%) in UKB controls (P<1x10^−5^). *SSPN* is located within DGC of myocytes and its activity protects against cardiac contractility impairment resulting from dystrophin deficiency in Duchenne muscular dystrophy, while loss of function destabilizes muscle adhesion and force generation^30,31^. Exploratory analysis of ultra-rare variants (MAF<1x10^−5^) in UKB highlighted additional associations with DCM (*SLC38A6* and *SSPN*) (**Table S13**).

To identify the organs, tissues and cell types mediating genetic risk of DCM, we performed bulk tissue-level heritability enrichment analysis. Cardiac and other muscle-related tissues (including vascular and gastrointestinal smooth muscle) were the most highly enriched (**Figure S8, Table S14**). Cell type heritability was assessed using the *sc-linker* framework^32^, by integrating single nuclei RNA-sequencing^33^ of LV tissue from 52 DCM cases with end-stage heart failure undergoing cardiac transplantation, and 18 controls, and genome-wide enhancer-promoter contact in the LV, with GWAS heritability. We highlight biologically important cell types and state-disease relationships by identifying enrichments in basal gene expression profiles within cardiomyocytes, and DCM-specific differentially expressed genes (DEG) in cardiomyocytes, fibroblasts, and mural cells (**Figure S8, Tables S15** and **S16**). When evaluating gene expression in control hearts, most prioritized genes had highest levels of expression in cardiomyocytes (**Figure S8**). Several of the prioritized DCM genes (notably including, *SSPN, MAP3K7*, and *NEDD4L*) were differentially expressed in cardiomyocytes in DCM (**Figure S8**). Supporting the important role of non-cardiomyocytes in DCM pathogenesis, fibroblasts and mural cells (primarily pericytes) consistently had the highest proportion of DEGs in enriched biological pathways (**Figure S8**), with most prioritized genes being DEGs in non-cardiomyocytes.

To explore non-cardiomyocyte and cardiomyocyte cell-non autonomous mechanisms and the role of prioritized genes encoding for ligands or receptors, we investigated intercellular signaling pathways using *CellChat*^*34*^, a method that uses cellular transcriptomics, a priori knowledge of ligand-receptor-cofactor interactions, and law of mass action to quantify communication networks. In DCM, there was an overall increase in global signaling, with notable reductions in cardiomyocyte-cardiomyocyte interaction strength (**Figure S9**). There was an increase in prioritized gene enriched TGF-β signaling pathway, and in pathways of specific prioritized genes (*COL4A1* and *EPHB1* interactions increased, and *THBS1* reduced) (**Figure S9**). While there were modest increases in overall collagen signaling in DCM, *COL4A1* expression was increased in fibroblasts in DCM (**Figure S8**) with increased signaling to cardiomyocytes, fibroblasts and mural cells (**Figure S8**). We demonstrate that *EPHB1* expression is highest in cardiomyocytes and its major ligand *EFNB2* is produced in endothelial cells, with an increase in ligand and corresponding decrease in receptor production in DCM (**Figure S8**). Similar findings were seen in a single nuclei study of pressure overloaded human hearts^35^. *BMPR1A* was predominantly expressed in cardiomyocytes, with increased expression in mural cells and fibroblasts leading to increased *BMP6*-*BMPR1A* signaling from endocardial cells to cardiomyocyte and fibroblast (**Figure S8**) as has previously been described^33^.

Given the importance of common genetic variation on DCM heritability, we generated a polygenic score (PGS_DCM_) with 4,309,853 SNP predictors and evaluated it in 347,585 unrelated participants of White British ancestry in UKB (**Figure 5A**). PGS was associated with DCM (OR per PGS SD 1.76 [95% CI 1.64 to 1.90], P <2x10^−16^; AUROC 0.71) in the general population, with 4-fold increased risk in the top centile compared with the median (OR 3.83, 95% CI 2.52 to 5.79, P 2.1x10^−10^) and 7-fold compared with the bottom centile (OR 7.04, 95% CI 2.42 to 20.52, P 3.5x10^−4^) (**Figures 5B** and **5C**). In 25,443 individuals with measured CMR traits, PGS_DCM_ was associated with a cardiac traits consistent with DCM (**Table S17**): reduced contractility (LVEF: per PGS SD –0.7%, P_adj_ 8.1x10^−78^; top vs. bottom centile 57.6 vs. 60.8, P_adj_ 1.7x10^−6^) and increased volumes (LVEDV: +2.1ml, P_adj_ 2.5x10^−45^; top vs. bottom centile: 158.1 vs. 143.4, P 3.1x10^−6^; LVESV: +1.9, P 1.6x10^−93^; top vs. bottom centile: 67.7 vs. 56.6, P 1.4x10^−9^). Given that penetrance and expressivity of DCM in carriers of rare pathogenic variants can be highly variable^36^, we next evaluated whether common variants affect penetrance of rare variants as has been demonstrated for hypertrophic cardiomyopathy^10^. In 1,546 carriers of pathogenic variants in DCM-causing genes in UKB (prevalence 0.5%), PGS_DCM_ stratified DCM prevalence (top quintile: 7.3%, bottom quintile: 1.7%, P 0.005), including in 1,166 carriers of rare *TTN* PTVs (**Figure 5D**). Finally, phenome-wide association study of PGS_DCM_ was used to explore genetic relationships between common variant risk and other traits, identifying significant associations with several heart failure and related cardiovascular phenotypes (electrophysiologic and valvular), and established risk factors for impaired cardiac function (hypertension and obesity) (**Figure 5E**). We also identified significant associations with cardiac ischemic phenotypes highlighting the shared genetic overlap between primary and ischemic heart failure, and inverse associations with hypertrophic cardiomyopathy that have recently been described^8^.

**Figure 5:**
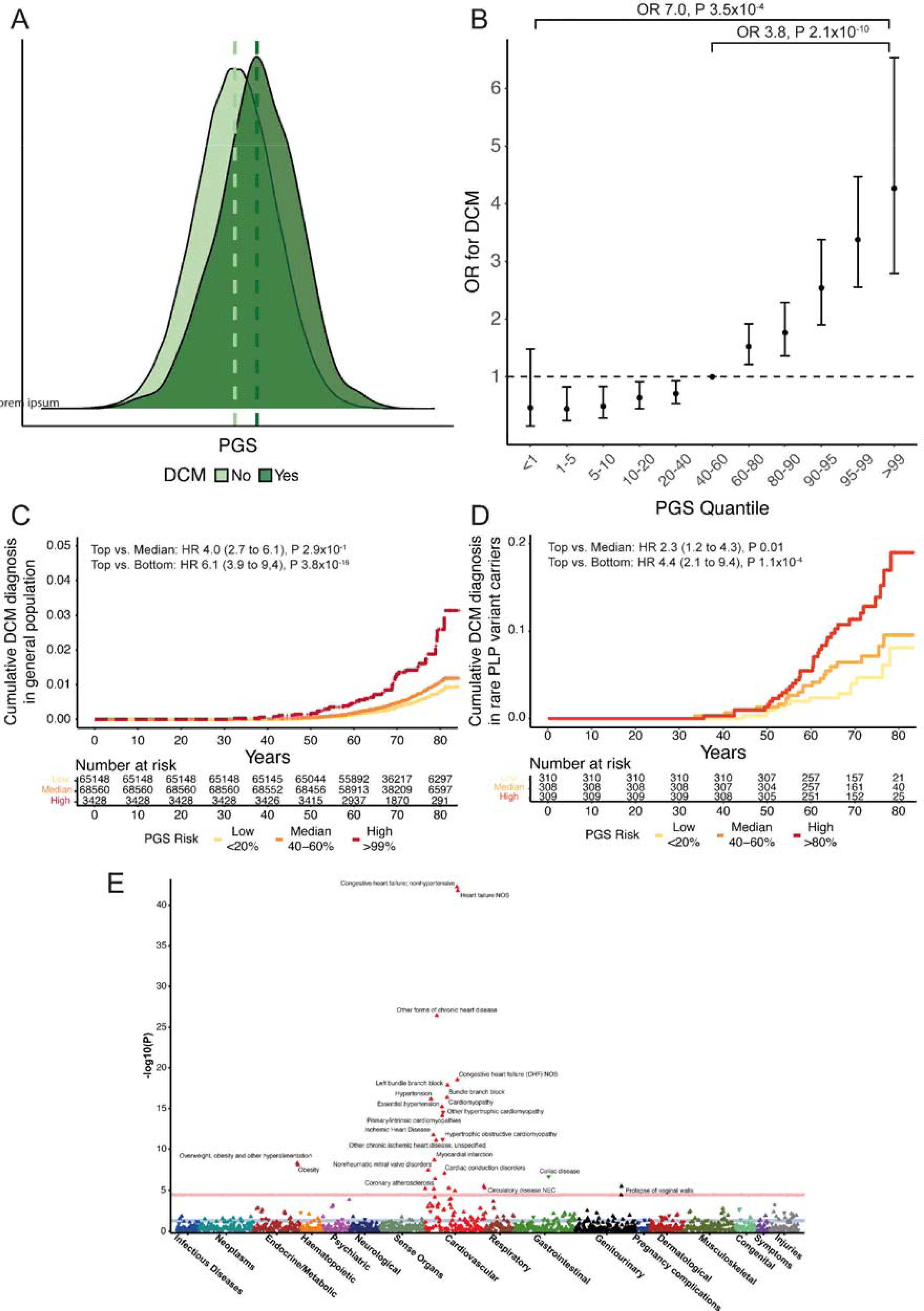
DCM PGS is associated with DCM disease status in the UK Biobank, including in carriers of pathogenic or likely pathogenic variants in DCM-causing genes. (A) PGS distribution in 347,585 UKB participants with and without DCM highlighting higher PGS in those with DCM. (B) Odds ratio for DCM in quantile bins compared with median (40-60%), demonstrating an increased risk of DCM in individuals with the highest PGS. (C) Cumulative hazards for lifetime diagnosis of DCM stratified by high (highest centile – red), median (middle quintile - orange) and low PGS (bottom quintile – yellow) in UKB. (D) Cumulative hazards for lifetime diagnosis of DCM in carriers of pathogenic or likely pathogenic (PLP) rare variants in DCM-causing genes, stratified by high (highest quintile – red), median (middle quintile - orange) and low PGS (bottom quintile – yellow) in UKB. (E) Manhattan plot of DCM PGS phenome-wide association study in UKB, showing associations with cardiovascular phenotypes and obesity. ICD-9 and ICD-10 diagnostic codes are mapped to Phecode Map version 1.2. Mapped phenotypes exceeding phenome-wide significance threshold (P 2.7x10^−5^, red line) are labelled. Blue line indicates nominal significance (P<0.05). Direction of triangle indicates the direction of effect of the PGS association.

In conclusion, we have performed the largest DCM GWAS to date, improving discovery power using MTAG with genetically correlated CMR traits, and identified 80 significant loci of which 66 have not been previously linked to DCM. Through a systematic locus annotation and gene prioritization approach, we identify effector genes at 61 loci, highlighting key biological pathways in disease pathogenesis. We demonstrate using single nuclei transcriptomics from explanted end-stage DCM hearts the importance of these biological pathways and the role of non-cardiomyocyte cell types and intercellular communication, including Ephrin-B and BMP6 signaling. Rare variant association testing identifies potential novel causes of DCM, including *MAP3K7, NEDD4L*, and *SSPN*. Finally, we generate a PGS that associates with DCM, and modulates penetrance of rare variants. These findings provide mechanistic insights into the genetic causes underlying DCM pathogenesis and may inform therapeutic strategies for DCM patients and at-risk individuals.

## Data Availability

Data from UK Biobank can be requested from the UK Biobank Access Management System. Data from 100,000 Genomes Project can be accessed following application to join the Genomics England Clinical Interpretation Partnership. GWAS summary statistics will be made available on the Cardiovascular Disease Knowledge Portal (https://cvd.hugeamp.org/) upon publication following peer-review. The PGS will be made available on the Polygenic Score Catalog (www.pgscatalog.org) following peer-review. The raw single nuclei gene expression dataset is available from CZI CELLxGENE (cellxgene.cziscience.com). The analyses reported in this article rely on previously published software, as detailed in the Methods.

## Acknowledgements and Funding Sources

This work was supported by funding from the British Heart Foundation [RE/18/4/34215, FS/IPBSRF/22/27059, FS/15/81/31817, FS/ICRF/21/26019, RG/19/6/34387, BC/F/21/220106, FS/18/65/34186, SP/19/1/34461, SP/17/11/32885]; the Medical Research Council [MC_UP_1605/13]; Wellcome Trust [107469/Z/15/Z]; the National Institute for Health Research (NIHR) Imperial College Biomedical Research Centre; NIHR Royal Brompton Cardiovascular Biomedical Research Unit; Sir Jules Thorn Charitable Trust [21JTA]; National Heart Lung Institute Foundation; Royston Centre for Cardiomyopathy Research; Rosetrees Trust. This research has been conducted in part using the UK Biobank Resource under Application Numbers 9922, 15422, 18545, 40616 and 47602.

The views expressed in this work are those of the authors and not necessarily those of the funders. For the purpose of open access, the authors have applied a Creative Commons Attribution (CC BY) licence to any Author Accepted Manuscript version arising from this submission.

## Competing interests

AH and RTL received funding from Pfizer Inc. J.S.W. have acted as a consultant for MyoKardia, Pfizer, Foresite Labs, and Health Lumen, and received institutional support from Bristol-Myers Squibb and Pfizer Inc. PC Personal fees for consultancies, outside the present work, for Amicus, Pfizer Inc, Owkin and Bristol Myers Squibb. M-P.D. declares holding equity in Dalcor Pharmaceuticals, unrelated to this work. The authors who are affiliated with deCODE genetics/Amgen Inc. and Regeneron Pharmaceuticals declare competing financial interests as employees. The remaining authors declare no competing interests.

## Online Methods

### Phenotype and study populations

Dilated cardiomyopathy (DCM) was defined in each participating study, using a harmonized, rule-based, multi-modal phenotyping algorithm as a guide. DCM was defined as left ventricular (LV) systolic dysfunction with or without LV dilatation in the absence of secondary causes of heart failure (coronary artery disease, valvular heart disease or congenital heart disease) (see **Supplementary Methods** for full definitions). Individuals with coronary artery disease, valvular heart disease or congenital heart disease were excluded from the control group. Imaging evidence or physician adjudication was preferred, but where unavailable, classifiers were defined as the presence of at least one relevant diagnosis or procedural code from the patient’s medical records.

### Discovery GWAS and multi-trait analysis of GWAS

The current genome-wide association study (GWAS) meta-analysis included 14,255 cases and 1,199,156 controls of European ancestry from 16 studies in the HERMES Consortium (cohorts described in **Supplementary Methods** and **Table S1**). Genotyping for 15 of 16 studies were performed locally in each participating study using high-density genotyping arrays imputed against reference whole-genome sequencing panels from the Haplotype Reference Consortium (HRC) (14 studies), 1000 Genomes Project (Garnier *et* al^37^), or population-specific reference panels (Estonian Biobank and deCODE) (**Supplementary Methods**). Genotyping for the Genomics England cohort was done using whole genome sequencing. Genetic association test was performed per study per phenotype, using a logistic regression model assuming additive genetic effects with adjustments for age, sex, genetic principal components (PC), and study-specific covariates. Full details of study-level GWAS methods available in **Supplementary Methods**. Descriptions of studies and participant characteristics are described in Table S1. Sensitivity analysis GWAS and meta-analysis of strictly defined DCM (**Supplementary Methods**) was performed using the same workflow. To assess the effects of ascertainment of DCM using strict criteria, GWAS meta-analysis was performed for the 6 studies that used strict criteria (**Supplementary Table 1**) and genetic correlation was assessed using bivariate linkage disequilibrium score regression with LDSC software^38^.

GWAS meta-analysis was performed centrally using METAL^39^ with an inverse-variance weighted fixed-effect model. To boost discovery power, we further conducted a multi-trait analysis of GWAS (MTAG), a method of jointly analyzing summary statistics from multiple overlapping GWAS of genetically correlated traits. GWAS in the UK Biobank of 10 cardiac MRI-derived LV traits (ejection fraction [LVEF], end-systolic volume [LVESV], end-diastolic volume [LVEDV], stroke volume, global circumferential, longitudinal and radial strains, mass, concentricity, and maximum wall thickness) from 36,083 unrelated participants of White British ancestry and without heart failure, cardiomyopathy, previous myocardial infarction, or structural heart disease^7^ were tested for genetic correlation with primary GWAS using *ldsc*^*40,41*^. MTAG of the primary GWAS was then performed with CMR traits with high genetic correlation (|r_g_| >0.7) using the *mtag* software^9^.

### SNP-based heritability estimation

The proportion of variance in HF risk explained by common SNPs, i.e. SNP-based heritability *(h*^*2*^_*SNP*_), was estimated from GWAS meta-analysis summary statistics using Linkage-Disequilibrium Adjusted Kinships (LDAK) SumHer software with the BLD-LDAK heritability model^6^. The *h*^*2*^_*SNP*_ estimates were calculated on a liability scale, which assumes that a binary phenotype has an underlying continuous liability, and above a certain liability threshold an individual becomes affected^42^. To model the expected heritability tagged by each SNP, we used pre-computed tagging files derived from 2,000 individuals White British individuals, and used a correction for sample prevalence by calculating the effective sample size assuming an equal number of cases and controls^43^. The conversion to liability scale was calculated using a population prevalence of 0.004 for DCM-Strict (based on estimated prevalence of 1 in 250 individuals^2,3^) and 0.08 for DCM (assuming twice the prevalence of DCM-Strict).

### Locus identification

To identify genetic susceptibility loci for DCM, we first identified conditionally independent genetic variants using a chromosome-wide stepwise conditional-joint analysis implemented in the Genome-wide Complex Trait Analysis (GCTA) software^44^ at a genome-wide significance threshold of *P<*5x10^−8^ in all GWAS, and additionally at FDR<1% estimated using *qvalue*) for DCM_GWAS_. To define a genomic locus, conditionally independent genetic variants across both DCM_GWAS_ and MTAG that are located within 500kb of each other were aggregated, and an additional 500kb were added to flank the variants at the extremes within each set. A genomic locus was considered novel if all conditionally independent variants within the locus are located ≥250kb away and not in LD (*R*^*2*^) with any sentinel variant with a P <5x10^−8^ reported in previously published GWAS of DCM^8,37^.

### Functionally-informed fine-mapping of genomic loci

To prioritize likely causal variants at each genomic locus, we performed functionally informed fine-mapping using PolyFun^45^ and SuSiE^46^. Using precomputed prior causal probabilities of 19 million imputed SNPs with a minor allele frequency (MAF)>0.001 based on meta-analysis of 15 traits in UK Biobank from PolyFun, we first estimate per-SNP heritability. These results were then passed to SuSiE to calculate per-SNP posterior inclusion probability (PIP) and to identify 95% credible sets of likely causal variants, assuming at most 5 causal variants per locus. To run fine-mapping, we used linkage disequilibrium (LD) reference panels from 10,000 randomly selected UK Biobank (UKB) European ancestry participants. The procedure was performed separately for locus identified from GWAS_DCM_ and GWAS_MTAG_ using the respective summary statistics. For each locus, variants within the identified 95% credible sets in either GWAS_DCM_ or GWAS_MTAG_ were aggregated, and annotated with nearest gene(s), genic functions, and Combined Annotation-Dependent Depletion (CADD) Phred score^47^ extracted from ANNOVAR^48^ and OpenTargets Genetics^49^.

#### Prioritization of effector genes at DCM loci

To systematically identify and prioritize effector genes at each locus, we followed a two-step approach. First, the nearest gene and the top 3 genes prioritized by either PoPS^50^ or V2G^51^ were selected as candidate genes. Second, the totality of evidence including nearest gene, PoPS, V2G and 5 additional approaches (coding variant, co-localization with gene expression, TWAS, ABC-model, and established Mendelian cardiomyopathy- and muscle-disease-causing genes) was summarized by identifying the number of individual approaches that identified each candidate gene as the most likely, assuming that it met each method’s minimum threshold for significance or relevance. Each method received equal weighting, with a maximum score of 8, and the candidate gene with the highest score at each genomic locus was determined to be the prioritized gene.

##### Transcriptome-wide association study

We estimated the associations between overall gene expression across tissues and DCM through a multi-tissue transcriptome-wide association study (TWAS) using expression quantitative trait loci (eQTL) data across 49 human tissues from GTEx v8 and the GWAS_DCM_ summary statistics implemented in S-MulTiXcan with MASH-R model^52^.

##### Colocalization with gene expression

To test the hypothesis whether genetic associations with gene expression in a given tissue and with DCM is driven by the same causal variants, we performed a statistical colocalization analysis using R *coloc* package^46^ allowing for multiple causal variants. The colocalization analysis was performed for all genes overlapping the identified DCM genetic loci using summary-level eQTL data from GTEx v8^53^ in tissue with a lowest TWAS *P-*value and the GWAS_DCM_ summary statistics.

##### Polygenic priority score

We computed the polygenic enrichment of gene features derived from cell-type specific gene expression, biological pathways, and protein-protein interactions for all protein-coding genes within the human genome using the Polygenic Priority Score (PoPS)^50^. A higher score implicates a higher probability of a gene being causal for the trait under study, given feature similarities to other predicted causal genes.

##### Variants-to-Gene

The variants-to-gene (V2G) model aggregates data from molecular phenotype quantitative trait loci experiments (eQTL, pQTL, and sQTL), chromatin interaction experiments, in silico functional predictions, and genomic distance (between the variant and a gene’s canonical transcriptional start site) to compute a variant-level score with a higher value reflecting a higher functional relevance on a given gene^51^. To map variant-level V2G score into gene-level score for gene prioritization, we extracted the V2G score for all variants that are in LD (*R*^*2*^ *> 0*.*8*) with conditionally independent variants or within the fine-mapped variant set for a given locus, and took the maximum V2G for a given gene.

##### Activity-by-Contact model

The Activity-by-Contact (ABC) model uses experimental estimates of enhancer activity (ATAC-seq, DNase-seq or H3K27ac ChIP-seq) and enhancer-promoter contact frequency (HiC) to predict enhancer-gene interactions^54^. Precomputed ABC scores generated from experimental data of cardiac left ventricles in ENCODE^55^ were identified for the genomic coordinates of fine mapped and lead variants, with scores >0.02 indicating an important interaction.

#### Rare variant gene-based association testing

Gene-based association testing was performed in the UK Biobank and 100,000 Genomes Project for all genes located within genomic loci, using the genome-wide regression test implemented by *REGENIE*. A whole genome regression model was fitted to allow handling of polygenicity, relatedness, and ancestry, using directly genotype-arrayed variants passing QC (MAF>0.01, <10% missingness, Hardy-Weinberg equilibrium [HWE] test P>10^−15^) in UKB, or directly sequenced variants in 100,000 Genomes Project (GeL). Next, gene-based burden test was performed conditional upon the phenotype-specific predictors from the genome-wide regression model and adjusting for sex, age, age^2^ and first ten genetic PCs, with body surface area and systolic blood pressure included as additional covariates for quantitative traits. The outcomes tested were binary case-control status (DCM [strict and broad definition], heart failure, and HCM) and in the UKB, related cardiac magnetic resonance imaging (CMR) quantitative traits (LVESV, LVEDV, LVEF, LVSV, and maximum LV wall thickness). Firth correction was applied to account for case-control imbalance. Burden tests collapse variants into a single variable that can be tested for association with a phenotype or trait, thereby reducing computational cost and test statistic inflation that is seen with other gene-based rare variant tests (for example, SKAT and SKAT-O). Individuals with missing phenotype data were dropped from analysis. For consistency across UKB and GeL, one rare variant mask of predicted truncating variants (start lost, stop gained, frameshift, splice acceptor or donor lost) with a MAF<1x10^−3^ was tested. To minimize the false positive rate resulting from genes with very low allele counts, a minimum allele count (MAC) threshold was applied which approximately considered the sample size: analysis in UKB required MAC≥20 for binary traits, and MAC≥3 for quantitative traits; and analysis in GeL required MAC≥3. P-values were adjusted for the total number of genes passing the MAC threshold that were tested. Exploratory results evaluating the effect of ultra rare (MAF<1x10^−5^) variants on binary outcomes in UKB were also tested.

To characterize the overall genetic architecture of DCM, gene-based burden testing was also performed for 16 DCM genes with moderate or definitive evidence^28^ in UKB to generate risk estimates for carriers of rare variants with DCM and heart failure. Two masks were tested: PTV and missense variants, both with MAF<1x10^−3^, with nominally significant associations highlighted in for visualization.

### Pathway enrichment analysis of effector genes, differentially expressed genes, and intercellular communication in DCM single-nuclei transcriptomics

Pathway gene ontology (GO) enrichment of effector genes and DEGs in DCM at cell type and state level, with identification of driver GO terms was performed using a two-stage algorithm implemented by g:Profiler^56^. For enrichment of DEGs, only highly expressed genes within a cell type or state were considered in the background. Driver GO terms were highlighted using a two-stage algorithm implemented by g:Profiler to identify pathways that were further examined in the DCM single nuclei dataset. DCM DEGs were required to have normalized log_2_ count >0.0125/nuclei and FDR<0.05. and differentially expressed genes (DEG) in DCM cell types.

To highlight the importance of cardiomyocyte and non-cardiomyocyte cell types in DCM, and the role of candidate genes and effector gene enriched signaling pathways, we explored disease-specific intercellular communication. The single nuclei transcriptome of DCM and control samples was interrogated using the *CellChat* package for manually curated ligand-receptor interactions (*CellChatDB*)^34^. In brief, this method identifies over-expressed genes within cell types and states, quantifies receptor-ligand communication probability between cells using the law of mass action, and infers statistically and biologically important cellular communications^34^. *CellChat* was run using default program settings, and analysed at the cell type level. Endocardial cells were separated out from other endothelial cells due to previously reported important biological effects on ligand-receptor signaling^33^.

### Tissue, cell type and cell state heritability enrichment

Tissue-level heritability enrichment analysis was performed using pre-calculated LD scores of gene expression data from GTEx^53^ and chromatin data from Roadmap Epigenomics^57^ and ENCODE^55^ projects, using *ldsc*^58^. For cell type and state heritability enrichment, we used the *sc-linker*^32^ approach to link transcriptome wide gene programs from single nuclei dataset with GWAS summary statistics. Gene programs derived from snRNA-seq were used to investigate heritability enrichment in cardiac cell types and states using the *sc-linker* framework^32^. This approach uses snRNA-seq data to generate gene programs that characterize individual cell types and states. These programs are then linked to genomic regions and the SNPs that regulate them by incorporating Roadmap Enhancer-Gene Linking^57,59^ and ABC models^54,60^. Finally, the disease informativeness of resulting SNP annotations is tested using stratified LD score regression (S-LDSC)^61^ conditional on broad sets of annotations from the baseline-LD model^38,62^.

Cell type and state-specific gene programs were generated from snRNA-seq data of ventricular tissue from 18 control subjects, with cell type and state annotations made as part of a larger study of ∼880,000 nuclei (samples from 52 DCM and 18 control subjects)^33^. Cell states that may not represent true biological states (for example, technical doublets) were excluded from analysis. For cell type and state disease-specific programs, pseudobulked counts were used to compare expression levels in DCM and control LV samples within all annotated cell types and cell states, implemented using *edgeR*^63^. Significant differentially expressed genes (DEG) were defined as FDR <0.05 and absolute(log_2_ fold change) >0.5, requiring a minimum normalized log_2_ count >0.0125 in both control and DCM samples.

### Polygenic risk score generation and testing

Polygenic score (PGS) was generated using a Bayesian framework that models ancestry-specific LD using an external reference set and uses a continuous shrinkage prior, implemented using the *PRS-CS* package^64^. The phi constant was automatically selected by PRS-CS in an unsupervised approach (*PRS-CS auto*). Whole genome PGS scores for all included UKB individuals were calculated using the PLINK 1.9 *--score* function^65^. Individual SNP weighted scores were generated from GWAS that excluded the UKB cohort, and a subsequent MTAG, to avoid the substantial inflation that is seen when there is overlap of individuals in the GWAS and testing cohorts^66^. The base GWAS and MTAG summary statistics were filtered to exclude rare and uncommon variants (MAF <0.01), and ambiguous SNPs that were not resolvable by strand flipping. We calculated a PGS for unrelated (3^rd^ degree or closer), White British participants in the UKB (application number 47602) using variants that passed genotyping QC (MAF>0.01, genotyping rate >0.99, HWE 1x10^−6^). Variants overlapping the base, target, and LD reference set (1000 Genomes Project Phase 3 European ancestry) were included. PGS predictive performance was assessed by area under the receiver operating characteristic, and association with DCM and associated CMR traits (OR per PGS standard deviation, and comparing top quantiles with the median) in the UKB, and in carriers of rare variants predicted to cause DCM^28^ (see **Supplementary Methods** for full details on variant curation and genes tested).

### Phenome-wide association study

The pleiotropic effect of genetic risk arising from common variants was tested by performing a phenome-wide association study (pheWAS) of PGS in the UKB. ICD-9 and ICD-10 codes from death records and hospital admission episodes were translated to Phecodes (Phecode Map 1.2)^67^. For binary phenotypes with at least 20 cases, PGS-phenotype association was tested using logistic regression adjusted for age, age^2^, sex and first ten genetic PCs as covariates. Significance threshold was adjusted for the total number of phenotypes tested (P<2.72x10^−5^), and data presented with Manhattan plots grouping by body system. PheWAS was performed using *PheWAS*^68^ in R version 4.0.3.

## Notes

### Competing Interest Statement

The authors have declared no competing interest.

