## Supplementary Method for "Genome-wide association analysis reveals insights into the molecular etiology underlying dilated cardiomyopathy"

**Supplementary Methods**

Study definitions for phenotyping of cases and controls

Individual study cohort descriptions

Study level GWAS methods and QC

Published dilated and non-ischaemic cardiomyopathy GWAS

Curation of pathogenic and likely pathogenic rare variants in DCM-causing genes in the UK Biobank

Individual study acknowledgements

HERMES Consortium Contributors

Supplementary Method References

### **Study definitions for phenotyping of cases and controls**

The primary GWAS phenotype of DCM required the presence of LV systolic dysfunction (LVSD) in the absence of coronary artery disease, valvular heart disease, or congenital heart disease (see below). Participants without heart failure, coronary artery disease ,valvular heart disease, or congenital heart disease were selected as controls.

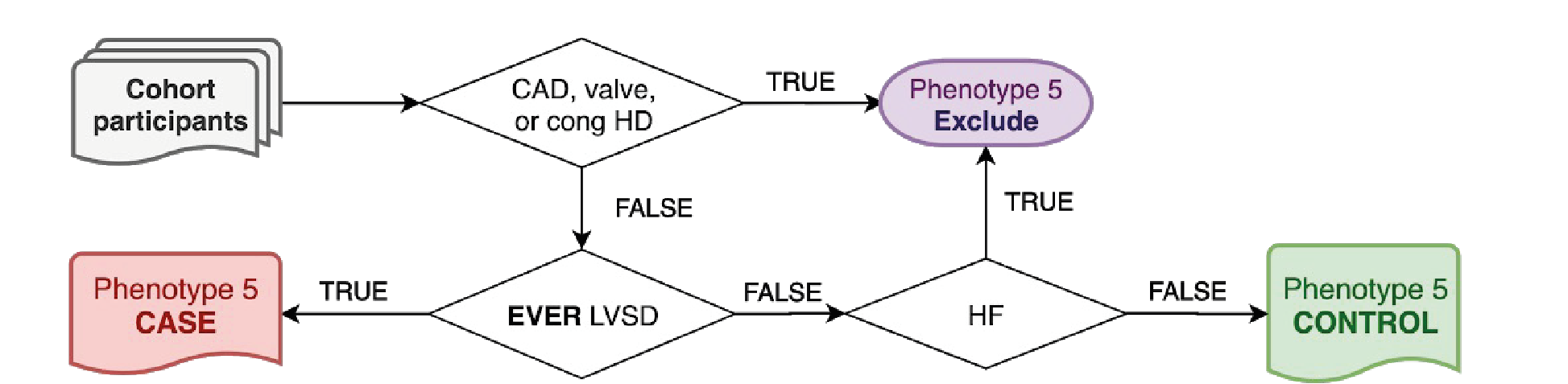

A stricter DCM diagnosis was used for sensitivity analysis and required a clinical diagnosis of DCM based on clinical guidelines, or documented ICD-10 code (I42.0) in the absence of coronary artery, valvular heart disease, or congenital heart disease codes (see below).

**LVSD:**

- LVEF <50% on cardiac imaging
- Manually adjudicated diagnosis of heart failure with reduced ejection fraction (HFrEF), heart failure with mid-range ejection fraction HFmrEF, systolic heart failure, DCM, permanent pacemaker implantation, arrhythmogenic cardiomyopathy, inflammatory cardiomyopathy and chemotherapy-related cardiomyopathy.
- >1 occurrence of the following codes:

| **Source** | **Code** | **Description** |
| --- | --- | --- |
| ICD10 | I42.0 | Dilated cardiomyopathy (Congestive cardiomyopathy) |
| ICD10 | I42.6 | Alcoholic cardiomyopathy |
| ICD10 | I42.7 | Cardiomyopathy due to drug and external agent |
| ICD10 | I25.5 | Ischaemic cardiomyopathy |
| ICD10 | I50.2 | Systolic (congestive) heart failure |
| ICD10 | O90.3 | Cardiomyopathy in the puerperium |
| ICD9 | 428.2 | Systolic heart failure |
| ICD9 | 674.5 | Peripartum cardiomyopathy |
| ICD9 | 425.5 | Alcoholic cardiomyopathy |
| ICD9 | 425.9 | Secondary cardiomyopathy, unspecified |
| OPCS4 | K61.7 | Cardiac resynchronisation device |
| CPT4 | 33224 | Insertion of pacing electrode, cardiac venous system, for left ventricular pacing, with attachment to previously placed pacemaker or implantable defibrillator pulse generator  (including revision of pocket, removal, insertion, and/or replacement of existing generator) |
| CPT4 | 33225 | Insertion of pacing electrode, cardiac venous system, for left ventricular pacing, at time of insertion of implantable defibrillator or pacemaker pulse generator (e.g., for upgrade to dual chamber system) (List separately in addition to code for primary procedure) |
| CPT4 | 33226 | Repositioning of previously implanted cardiac venous system (left ventricular) electrode (including removal, insertion and/or replacement of existing generator) |

**Coronary artery disease (CAD), valvular heart disease (VHD) or congenital heart disease (CHD):**

- Manually adjudicated diagnosis of myocardial infarction or revascularization.
- Severe primary valve disease defined by a relevant surgical or percutaneous valve procedure.
- Any congenital malformation of the heart or great vessels defined by diagnosis or relevant surgical or percutaneous procedure.
- Any rheumatic heart valve disease defined by diagnosis.
- >1 occurrence of the following codes:

| **Source** | **Code** | **Description** | **Phenotype class** |
| --- | --- | --- | --- |
| ICD10 | I21.* | Acute myocardial infarction | CAD |
| ICD10 | I22.* | Subsequent myocardial infarction | CAD |
| ICD10 | I23.* | Certain current complications following acute myocardial infarction | CAD |
| ICD10 | I24.1 | Dressler's syndrome | CAD |
| ICD10 | I25.2 | Old myocardial infarction | CAD |
| ICD10 | I25.5 | Ischaemic cardiomyopathy | CAD |
| ICD10 | I25.6 | Silent myocardial ischaemia | CAD |
| ICD10 | 745.* | Bulbus cordis anomalies and anomalies of cardiac septal closure | CHD |
| ICD10 | Q20.* | Congenital malformations of cardiac chambers and connections | CHD |
| ICD10 | Q21.* | Congenital malformations of cardiac septa | CHD |
| ICD10 | Q22.* | Congenital malformations of pulmonary and tricuspid valves | CHD |
| ICD10 | Q23.* | Congenital malformations of aortic and mitral valves | CHD |
| ICD10 | Q24.* | Other congenital malformations of heart | CHD |
| ICD10 | Q25.* | Congenital malformations of great arteries | CHD |
| ICD10 | Q26.* | Congenital malformations of great veins | CHD |
| ICD10 | 746.* | Other congenital anomalies of heart | VHD |
| ICD10 | I05.* | Rheumatic mitral valve diseases | VHD |
| ICD10 | I06.* | Rheumatic aortic valve diseases | VHD |
| ICD10 | I07.* | Rheumatic tricuspid valve diseases | VHD |
| ICD9 | 410.* | Myocardial infarction | CAD |
| ICD9 | 412.* | Old myocardial infarction | CAD |
| ICD9 | 394.* | Diseases of mitral valve | VHD |
| ICD9 | 395.* | Diseases of aortic valve | VHD |
| ICD9 | 396.* | Diseases of mitral and aortic valves | VHD |
| ICD9 | 397.* | Diseases of other endocardial structures | VHD |
| OPCS4 | K40.* | Saphenous vein graft replacement of coronary artery | CAD |
| OPCS4 | K41.* | Other autograft replacement of coronary artery | CAD |
| OPCS4 | K42.* | Allograft replacement of coronary artery | CAD |
| OPCS4 | K43.* | Prosthetic replacement of coronary artery | CAD |
| OPCS4 | K44.* | Other replacement of coronary artery | CAD |
| OPCS4 | K45.* | Connection of thoracic artery to coronary artery | CAD |
| OPCS4 | K46.* | Other bypass of coronary artery | CAD |
| OPCS4 | K47.* | Repair of coronary artery | CAD |
| OPCS4 | K48.* | Other open operations on coronary artery | CAD |
| OPCS4 | K49.* | Transluminal balloon angioplasty of coronary artery | CAD |
| OPCS4 | K50.* | Other therapeutic transluminal operations on coronary artery | CAD |
| OPCS4 | K75.* | Percutaneous transluminal balloon angioplasty and insertion of stent into coronary artery | CAD |
| OPCS4 | K25.* | Plastic repair of mitral valve | VHD |
| OPCS4 | K26.* | Plastic repair of aortic valve | VHD |
| OPCS4 | K27.* | Plastic repair of tricuspid valve | VHD |
| OPCS4 | K28.* | Plastic repair of pulmonary valve | VHD |
| OPCS4 | K29.* | Plastic repair of unspecified valve of heart | VHD |
| OPCS4 | K30.* | Revision of plastic repair of valve of heart | VHD |
| OPCS4 | K31.* | Open incision of valve of heart | VHD |
| OPCS4 | K32.* | Closed incision of valve of heart | VHD |
| OPCS4 | K34.* | Other open operations on valve of heart | VHD |
| OPCS4 | K36.* | Excision of valve of heart | VHD |
| OPCS4 | K37.* | Removal of obstruction from structure adjacent to valve of heart | VHD |
| OPCS4 | K38.* | Other operations on structure adjacent to valve of heart | VHD |
| CPT4 | 33510 | Coronary artery bypass, vein only; single coronary venous graft | CAD |
| CPT4 | 33510 | Coronary artery bypass, vein only; single coronary venous graft | CAD |
| CPT4 | 33511 | Coronary artery bypass, vein only; 2 coronary venous grafts | CAD |
| CPT4 | 33511 | Coronary artery bypass, vein only; 2 coronary venous grafts | CAD |
| CPT4 | 33512 | Coronary artery bypass, vein only; 3 coronary venous grafts | CAD |
| CPT4 | 33512 | Coronary artery bypass, vein only; 3 coronary venous grafts | CAD |
| CPT4 | 33513 | Coronary artery bypass, vein only; 4 coronary venous grafts | CAD |
| CPT4 | 33513 | Coronary artery bypass, vein only; 4 coronary venous grafts | CAD |
| CPT4 | 33530 | Reoperation, coronary artery bypass procedure or valve procedure, more than 1 month after original operation (List separately in addition to code for primary procedure) | CAD |
| CPT4 | 33533 | Coronary artery bypass, using arterial graft(s); single arterial graft | CAD |
| CPT4 | 33533 | Coronary artery bypass, using arterial graft(s); single arterial graft | CAD |
| CPT4 | 33534 | Coronary artery bypass, using arterial graft(s); 2 coronary arterial grafts | CAD |
| CPT4 | 33534 | Coronary artery bypass, using arterial graft(s); 2 coronary arterial grafts | CAD |
| CPT4 | 33535 | Coronary artery bypass, using arterial graft(s); 3 coronary arterial grafts | CAD |
| CPT4 | 33535 | Coronary artery bypass, using arterial graft(s); 3 coronary arterial grafts | CAD |
| CPT4 | 33536 | Coronary artery bypass, using arterial graft(s); 4 or more coronary arterial grafts | CAD |
| CPT4 | 33536 | Coronary artery bypass, using arterial graft(s); 4 or more coronary arterial grafts | CAD |
| CPT4 | 92920 | Percutaneous transluminal coronary angioplasty; single major coronary artery or branch | CAD |
| CPT4 | 92920 | Percutaneous transluminal coronary angioplasty; single major coronary artery or branch | CAD |
| CPT4 | 92921 | Percutaneous transluminal coronary angioplasty; each additional branch of a major coronary artery | CAD |
| CPT4 | 92921 | Percutaneous transluminal coronary angioplasty; each additional branch of a major coronary artery | CAD |
| CPT4 | 92924 | Percutaneous transluminal coronary atherectomy, with coronary angioplasty when performed; single major coronary artery or branch | CAD |
| CPT4 | 92924 | Percutaneous transluminal coronary atherectomy, with coronary angioplasty when performed; single major coronary artery or branch | CAD |
| CPT4 | 92925 | Percutaneous transluminal coronary atherectomy, with coronary angioplasty when performed; each additional branch of a major coronary artery | CAD |
| CPT4 | 92925 | Percutaneous transluminal coronary atherectomy, with coronary angioplasty when performed; each additional branch of a major coronary artery | CAD |
| CPT4 | 92928 | Percutaneous transcatheter placement of intracoronary stent(s), with coronary angioplasty when performed; single major coronary artery or branch | CAD |
| CPT4 | 92928 | Percutaneous transcatheter placement of intracoronary stent(s), with coronary angioplasty when performed; single major coronary artery or branch | CAD |
| CPT4 | 92929 | Percutaneous transcatheter placement of intracoronary stent(s), with coronary angioplasty when performed; each additional branch of a major coronary artery | CAD |
| CPT4 | 92929 | Percutaneous transcatheter placement of intracoronary stent(s), with coronary angioplasty when performed; each additional branch of a major coronary artery | CAD |
| CPT4 | 92933 | Percutaneous transluminal coronary atherectomy, with intracoronary stent, with coronary angioplasty when performed; single major coronary artery or branch | CAD |
| CPT4 | 92933 | Percutaneous transluminal coronary atherectomy, with intracoronary stent, with coronary angioplasty when performed; single major coronary artery or branch | CAD |
| CPT4 | 92934 | Percutaneous transluminal coronary atherectomy, with intracoronary stent, with coronary angioplasty when performed; each additional branch of a major coronary artery | CAD |
| CPT4 | 92934 | Percutaneous transluminal coronary atherectomy, with intracoronary stent, with coronary angioplasty when performed; each additional branch of a major coronary artery | CAD |
| CPT4 | 92937 | Percutaneous transluminal revascularization of or through coronary artery bypass graft (internal mammary, free arterial, venous), any combination of intracoronary stent, atherectomy and angioplasty, including distal protection when performed; single vessel | CAD |
| CPT4 | 92937 | Percutaneous transluminal revascularization of or through coronary artery bypass graft (internal mammary, free arterial, venous), any combination of intracoronary stent, atherectomy and angioplasty, including distal protection when performed; single vessel | CAD |
| CPT4 | 92938 | Percutaneous transluminal revascularization of or through coronary artery bypass graft (internal mammary, free arterial, venous), any combination of intracoronary stent, atherectomy and angioplasty, including distal protection when performed; each additional branch subtended by the bypass graft | CAD |
| CPT4 | 92938 | Percutaneous transluminal revascularization of or through coronary artery bypass graft (internal mammary, free arterial, venous), any combination of intracoronary stent, atherectomy and angioplasty, including distal protection when performed; each additional branch subtended by the bypass graft | CAD |
| CPT4 | 92941 | Percutaneous transluminal revascularization of acute total/subtotal occlusion during acute myocardial infarction, coronary artery or coronary artery bypass graft, any combination of intracoronary stent, atherectomy and angioplasty, including aspiration thrombectomy when performed, single vessel | CAD |
| CPT4 | 92941 | Percutaneous transluminal revascularization of acute total/subtotal occlusion during acute myocardial infarction, coronary artery or coronary artery bypass graft, any combination of intracoronary stent, atherectomy and angioplasty, including aspiration thrombectomy when performed, single vessel | CAD |
| CPT4 | 92943 | Percutaneous transluminal revascularization of chronic total occlusion, coronary artery, coronary artery branch, or coronary artery bypass graft, any combination of intracoronary stent, atherectomy and angioplasty; single vessel | CAD |
| CPT4 | 92943 | Percutaneous transluminal revascularization of chronic total occlusion, coronary artery, coronary artery branch, or coronary artery bypass graft, any combination of intracoronary stent, atherectomy and angioplasty; single vessel | CAD |
| CPT4 | 92944 | Percutaneous transluminal revascularization of chronic total occlusion, coronary artery, coronary artery branch, or coronary artery bypass graft, any combination of intracoronary stent, atherectomy and angioplasty; each additional coronary artery, coronary artery branch, or bypass graft | CAD |
| CPT4 | 92944 | Percutaneous transluminal revascularization of chronic total occlusion, coronary artery, coronary artery branch, or coronary artery bypass graft, any combination of intracoronary stent, atherectomy and angioplasty; each additional coronary artery, coronary artery branch, or bypass graft | CAD |
| CPT4 | 92973 | Percutaneous transluminal coronary thrombectomy, mechanical* | CAD |
| CPT4 | 92973 | Percutaneous transluminal coronary thrombectomy, mechanical* | CAD |
| CPT4 | 92975 | Thrombolysis, coronary; by intracoronary infusion, including selective coronary angiography | CAD |
| CPT4 | 92975 | Thrombolysis, coronary; by intracoronary infusion, including selective coronary angiography | CAD |
| CPT4 | 33608 | Repair of complex cardiac anomaly other than pulmonary atresia with ventricular septal defect by construction or replacement of conduit from right or left ventricle to pulmonary artery | CHD |
| CPT4 | 33610 | Repair of complex cardiac anomalies (eg, single ventricle with subaortic obstruction) by surgical enlargement of ventricular septal defect | CHD |
| CPT4 | 33611 | Repair of double outlet right ventricle with intraventricular tunnel repair; | CHD |
| CPT4 | 33612 | Repair of double outlet right ventricle with intraventricular tunnel repair; with repair of right ventricular outflow tract obstruction | CHD |
| CPT4 | 33615 | Repair of complex cardiac anomalies (eg, tricuspid atresia) by closure of atrial septal defect and anastomosis of atria or vena cava to pulmonary artery (simple Fontan procedure) | CHD |
| CPT4 | 33617 | Repair of complex cardiac anomalies (e.g., single ventricle by modified Fontan) | CHD |
| CPT4 | 33619 | Repair of single ventricle with aortic outflow obstruction and aortic arch hypoplasia (hypoplastic left heart syndrome) (eg, Norwood procedure) | CHD |
| CPT4 | 33641 | Repair atrial septal defect, secundum, with cardiopulmonary bypass, with or without patch | CHD |
| CPT4 | 33645 | Direct or patch closure, sinus venosus, with or without anomalous pulmonary venous drainage | CHD |
| CPT4 | 33647 | Repair of atrial septal defect and ventricular septal defect, with direct or patch closure | CHD |
| CPT4 | 33660 | Repair of incomplete or partial atrioventricular canal (ostium primum atrial septal defect), with or without atrioventricular valve repair | CHD |
| CPT4 | 33665 | Repair of intermediate or transitional atrioventricular canal, with or without atrioventricular valve repair | CHD |
| CPT4 | 33670 | Repair of complete atrioventricular canal, with or without prosthetic valve | CHD |
| CPT4 | 33675 | Closure of multiple ventricular septal defects; | CHD |
| CPT4 | 33676 | Closure of multiple ventricular septal defects; with pulmonary valvotomy or infundibular resection (acyanotic) | CHD |
| CPT4 | 33677 | Closure of multiple ventricular septal defects; with removal of pulmonary artery band, with or without gusset | CHD |
| CPT4 | 33681 | Closure of single ventricular septal defect, with or without patch; | CHD |
| CPT4 | 33684 | Closure of single ventricular septal defect, with or without patch; with pulmonary valvotomy or infundibular resection (acyanotic) | CHD |
| CPT4 | 33688 | Closure of single ventricular septal defect, with or without patch; with removal of pulmonary artery band, with or without gusset | CHD |
| CPT4 | 33692 | Complete repair tetralogy of Fallot without pulmonary atresia; | CHD |
| CPT4 | 33694 | Complete repair tetralogy of Fallot without pulmonary atresia; with transannular patch | CHD |
| CPT4 | 33697 | Complete repair tetralogy of Fallot with pulmonary atresia including construction of conduit from right ventricle to pulmonary artery and closure of ventricular septal defect | CHD |
| CPT4 | 33702 | Repair sinus of Valsalva fistula, with cardiopulmonary bypass; | CHD |
| CPT4 | 33710 | Repair sinus of Valsalva fistula, with cardiopulmonary bypass; with repair of ventricular septal defect | CHD |
| CPT4 | 33720 | Repair sinus of Valsalva aneurysm, with cardiopulmonary bypass | CHD |
| CPT4 | 33722 | Closure of aortico-left ventricular tunnel | CHD |
| CPT4 | 33732 | Repair of cor triatriatum or supravalvular mitral ring by resection of left atrial membrane | CHD |
| CPT4 | 33735 | Atrial septectomy or septostomy; closed heart (Blalock-Hanlon type operation) | CHD |
| CPT4 | 33736 | Atrial septectomy or septostomy; open heart with cardiopulmonary bypass | CHD |
| CPT4 | 33737 | Atrial septectomy or septostomy; open heart, with inflow occlusion | CHD |
| CPT4 | 33770 | Repair of transposition of the great arteries with ventricular septal defect and subpulmonary stenosis; without surgical enlargement of ventricular septal defect | CHD |
| CPT4 | 33774 | Repair of transposition of the great arteries, atrial baffle procedure (eg, Mustard or Senning type) with cardiopulmonary bypass; | CHD |
| CPT4 | 33776 | Repair of transposition of the great arteries, atrial baffle procedure (eg, Mustard or Senning type) with cardiopulmonary bypass; with closure of ventricular septal defect | CHD |
| CPT4 | 33780 | Repair of transposition of the great arteries, aortic pulmonary artery reconstruction (eg, Jatene type); with closure of ventricular septal defect | CHD |
| CPT4 | 33782 | Aortic root translocation with ventricular septal defect and pulmonary stenosis repair (ie,  Nikaidoh procedure); without coronary ostium reimplantation | CHD |
| CPT4 | 33783 | Aortic root translocation with ventricular septal defect and pulmonary stenosis repair (ie,  Nikaidoh procedure); with reimplantation of 1 or both coronary ostia | CHD |
| CPT4 | 33786 | Total repair, truncus arteriosus (Rastelli type operation) | CHD |
| CPT4 | 33813 | Obliteration of aortopulmonary septal defect; without cardiopulmonary bypass | CHD |
| CPT4 | 33814 | Obliteration of aortopulmonary septal defect; with cardiopulmonary bypass | CHD |
| CPT4 | 33920 | Repair of pulmonary atresia with ventricular septal defect, by construction or replacement of conduit from right or left ventricle to pulmonary artery | CHD |
| CPT4 | 33361 | Transcatheter aortic valve replacement (TAVR/TAVI) with prosthetic valve; percutaneous femoral artery approach | VHD |
| CPT4 | 33362 | Transcatheter aortic valve replacement (TAVR/TAVI) with prosthetic valve; open femoral artery approach | VHD |
| CPT4 | 33363 | Transcatheter aortic valve replacement (TAVR/TAVI) with prosthetic valve; open axillary artery approach | VHD |
| CPT4 | 33364 | Transcatheter aortic valve replacement (TAVR/TAVI) with prosthetic valve; open iliac artery approach | VHD |
| CPT4 | 33365 | Transcatheter aortic valve replacement (TAVR/TAVI) with prosthetic valve; transaortic approach (e.g., median sternotomy, mediastinotomy) | VHD |
| CPT4 | 33366 | Transcatheter aortic valve replacement (TAVR/TAVI) with prosthetic valve; transapical exposure  (e.g., left thoracotomy) | VHD |
| CPT4 | 33367 | Transcatheter aortic valve replacement (TAVR/TAVI) with prosthetic valve; cardiopulmonary bypass support with percutaneous peripheral arterial and venous cannulation (e.g., femoral vessels) (List separately in addition to code for primary procedure) | VHD |
| CPT4 | 33368 | Transcatheter aortic valve replacement (TAVR/TAVI) with prosthetic valve; cardiopulmonary bypass support with open peripheral arterial and venous cannulation (e.g., femoral, iliac, axillary vessels) (List separately in addition to code for primary procedure) | VHD |
| CPT4 | 33369 | Transcatheter aortic valve replacement (TAVR/TAVI) with prosthetic valve; cardiopulmonary bypass support with central arterial and venous cannulation (e.g., aorta, right atrium, pulmonary artery) (List separately in addition to code for primary procedure) | VHD |
| CPT4 | 33390 | Valvuloplasty, aortic valve, open, with cardiopulmonary bypass; simple (ie, valvotomy, debridement, debulking and/or simple commissural resuspension) | VHD |
| CPT4 | 33391 | Valvuloplasty, aortic valve, open, with cardiopulmonary bypass; complex (eg, leaflet extension, leaflet resection, leaflet reconstruction or annuloplasty) | VHD |
| CPT4 | 33400 | Aortic valvuloplasty | VHD |
| CPT4 | 33401 | Open valvuloplasty of aortic valve with inflow occlusion | VHD |
| CPT4 | 33403 | Valvuloplasty, aortic valve; using transventricular dilation, with cardiopulmonary bypass | VHD |
| CPT4 | 33404 | LV—aorta conduit | VHD |
| CPT4 | 33405 | Replacement, aortic valve, with cardiopulmonary bypass; with prosthetic valve other than homograft or stentless valve | VHD |
| CPT4 | 33406 | Replacement, aortic valve, with cardiopulmonary bypass; with allograft valve (freehand) | VHD |
| CPT4 | 33410 | Replacement, aortic valve, with cardiopulmonary bypass; with stentless tissue valve | VHD |
| CPT4 | 33411 | Replacement, aortic valve; with aortic annulus enlargement, noncoronary sinus | VHD |
| CPT4 | 33412 | Replacement, aortic valve; with transventricular aortic annulus enlargement (Konno procedure) | VHD |
| CPT4 | 33413 | Replacement, aortic valve; by translocation of autologous pulmonary valve with allograft replacement of pulmonary valve (Ross procedure) | VHD |
| CPT4 | 33415 | Resection or incision of subvalvular tissue for discrete subvalvular aortic stenosis | VHD |
| CPT4 | 33418 | Transcatheter mitral valve repair, percutaneous approach, including transseptal | VHD |
| CPT4 | 33419 | Transcatheter mitral valve repair, percutaneous approach, including transseptal puncture when performed; additional prosthesis(es) during same session (List separately in addition to code for primary procedure) | VHD |
| CPT4 | 33420 | Valvotomy, mitral valve; closed heart | VHD |
| CPT4 | 33422 | Valvotomy, mitral valve; open heart, with cardiopulmonary bypass | VHD |
| CPT4 | 33425 | Valvuloplasty, mitral valve, with cardiopulmonary bypass | VHD |
| CPT4 | 33426 | Valvuloplasty, mitral valve, with cardiopulmonary bypass; with prosthetic ring | VHD |
| CPT4 | 33427 | Valvuloplasty, mitral valve, with cardiopulmonary bypass; radical reconstruction, with or without ring | VHD |
| CPT4 | 33430 | Replacement, mitral valve, with cardiopulmonary bypass | VHD |
| CPT4 | 33440 | Replacement of aortic valve by translocation of autologous pulmonary valve and transventricular aortic annulus enlargement of left ventricular outflow tract with valved conduit replacement of pulmonary valve | VHD |
| CPT4 | 33460 | Valvectomy, tricuspid valve, with cardiopulmonary bypass | VHD |
| CPT4 | 33463 | Valvuloplasty, tricuspid valve; without ring insertion | VHD |
| CPT4 | 33464 | Valvuloplasty, tricuspid valve; with ring insertion | VHD |
| CPT4 | 33465 | Replacement, tricuspid valve, with cardiopulmonary bypass | VHD |
| CPT4 | 33468 | Tricuspid valve repositioning and plication for Ebstein anomaly | VHD |
| CPT4 | 33470 | Valvotomy, pulmonary valve, closed heart; transventricular | VHD |
| CPT4 | 33471 | Valvotomy, pulmonary valve, closed heart; via pulmonary artery | VHD |
| CPT4 | 33472 | Incision of valve at right lower heart chamber, open procedure | VHD |
| CPT4 | 33474 | Valvotomy, pulmonary valve, open heart, with cardiopulmonary bypass | VHD |
| CPT4 | 33475 | Replacement, pulmonary valve | VHD |
| CPT4 | 33476 | Right ventricular resection for infundibular stenosis, with or without commissurotomy | VHD |
| CPT4 | 33477 | Transcatheter pulmonary valve implantation, percutaneous approach, including prestenting of the valve delivery site, when performed | VHD |
| CPT4 | 33496 | Repair of non-structural prosthetic valve dysfunction with cardiopulmonary bypass (separate procedure) | VHD |
| CPT4 | 33600 | Closure of atrioventricular valve (mitral or tricuspid) by suture or patch | VHD |
| CPT4 | 33602 | Closure of semilunar valve (aortic or pulmonary) by suture or patch | VHD |
| CPT4 | 92986 | Percutaneous balloon valvuloplasty; aortic valve | VHD |
| CPT4 | 92987 | Percutaneous balloon valvuloplasty; mitral valve | VHD |
| CPT4 | 92990 | Percutaneous balloon valvuloplasty; pulmonary valve | VHD |
| CPT4 | 93355 | Echocardiography, transesophageal (TEE) for guidance of a transcatheter intracardiac or great vessel(s) structural intervention(s) (eg, TAVR, transcatheter pulmonary valve replacement, mitral valve repair, paravalvular regurgitation repair, left atrial ap | VHD |
| CPT4 | 93590 | Percutaneous transcatheter closure of paravalvular leak; initial occlusion device, mitral valve | VHD |
| CPT4 | 93592 | Percutaneous transcatheter closure of paravalvular leak; each additional occlusion device (List separately in addition to code for primary procedure) | VHD |
| CPT4 | 0256T | Implantation of catheter-delivered prosthetic aortic heart valve; endovascular approach | VHD |
| CPT4 | 0257T | Implantation of catheter-delivered prosthetic aortic heart valve; open thoracic approach (eg, transapical, transventricular) | VHD |
| CPT4 | 0262T | Implantation of catheter-delivered prosthetic pulmonary valve, endovascular approach | VHD |
| CPT4 | 0318T | Implantation of catheter-delivered prosthetic aortic heart valve, open thoracic approach, (eg, transapical, other than transaortic) | VHD |
| CPT4 | 0343T | Transcatheter mitral valve repair percutaneous approach including transseptal puncture when performed; initial prosthesis | VHD |
| CPT4 | 0345T | Transcatheter mitral valve repair percutaneous approach via the coronary sinus | VHD |
| CPT4 | 0483T | Transcatheter mitral valve implantation/replacement (TMVI) with prosthetic valve; percutaneous approach, including transseptal puncture, when performed | VHD |
| CPT4 | 0484T | Transcatheter mitral valve implantation/replacement (TMVI) with prosthetic valve; transthoracic exposure (e.g., thoracotomy, transapical) | VHD |

**Individual study cohort descriptions**

Full details on individual study genotyping is reported in the HERMES 2.0 study.

**Bio-SHiFT-TRIUMPH^1,2^ and Deotinchem Cohort Study**

Bio-SHiFT (Serial Biomarker Measurements and New Echocardiographic Techniques in chronic Heart Failure Patients Result in Tailored Prediction of Prognosis) is a prospective, observational study of stable patients with chronic heart failure (CHF). The study was conducted in Erasmus MC, Rotterdam, the Netherlands, and Noordwest Ziekenhuisgroep, Alkmaar, the Netherlands. Patients were recruited during their regular outpatient visits and were in clinically stable condition. Patients were eligible if aged ≥18 years and CHF was diagnosed ≥3 months ago according to the guidelines of the European Society of Cardiology. Both patients suffering from HF with reduced ejection fraction and HF with preserved ejection fraction were included. This study was approved by the medical ethics committee of the Erasmus MC, Rotterdam, the Netherlands, and was conducted in accordance with the Declaration of Helsinki. Written informed consent was obtained from all patients. The study is registered in ClinicalTrials.gov, number NCT01851538.
TRIUMPH (TRanslational Initiative on Unique and novel strategies for Management of Patients with Heart failure, NTR1893), is an observational prospective study of patients admitted with acute HF in 14 hospitals in the Netherlands between September 2009 and December 2013. Patients were eligible if aged ≥18 years and if they were hospitalized with decompensation of known chronic HF or newly diagnosed HF. The three other inclusion criteria were: 1) natriuretic peptide levels ≥3 times higher than the upper limit of normal; 2) evidence of sustained systolic or diastolic left ventricular dysfunction; and 3) treatment with intravenous diuretics. Patients with HF that was precipitated by a noncardiac condition, by severe valvular dysfunction without sustained left ventricular dysfunction, or by an acute ST-segment elevation myocardial infarction were excluded. Furthermore, patients scheduled for a coronary revascularization procedure, on a waiting list for heart transplantation, with severe renal failure for which dialysis was needed, or with a coexisting condition with a life expectancy <1 year could not participate. The study was approved by the medical ethics committees of all participating centers. All study participants provided written informed consent.

The Doetinchem Cohort Study is an ongoing prospective study in which participants are re-examined every 5 years. A general population sample of 7,769 males and females aged 20 to 59 years was examined in 1987-91 (first round) in Doetinchem, a town in the eastern part of the Netherlands and re-examined in a second (1993-1997), third (1998-2002), fourth (2003-2007), fifth (2008-2012), and sixth round (2013-2017). This study was approved according to the guidelines of the Helsinki Declaration by the external Medical Ethics Committee of the Netherlands Organization for Applied Scientific Research (first four rounds) and the Medical Ethics Committee of the University of Utrecht (later rounds). All participants provided written informed consent.

**BioVU**

The BioVU cohort consisted of patients at Vanderbilt University Medical Center who have consented to be a part of the program. Study cohort was derived from EHR data available via VUMC's Synthetic Derivative using the phenotype classifier described in the HERMES Consortium analysis plan. The Synthetic Derivative is the de-identified version of the VUMC EHR and similar to BioVU, contains DNA and genotype data from clinical blood samples that would otherwise be discarded. The Synthetic Derivative can be used in conjunction with BioVU to identify record sets for genome-phenome analysis.

**The Copenhagen Hospital Biobank Cardiovascular Study and The Danish Blood Donor Study (CHB)^3^**

This study is part of the study: "Genetics of cardiovascular disease” – a genome-wide association study on repository samples from Copenhagen Hospital Biobank - the CHB-CVS study. CHB has previously been described. CHB-CVS is a genetic study on patients from CHB with cardiovascular disease identified through the Danish National Patient Registry (NPR). CHB-CVS is approved by the National Committee on Health Research Ethics (NVC 1708829) and the Danish Data Protection Agency (P-2019-93). DBDS is a prospective cohort and biobank study on general health among blood donors. The DBDS has previously been described in detail. The genetic studies within DBDS were approved by the National Committee on Health Research Ethics (NVC 1700407) and the Data Protection Agency (P-2019-99). Cases and controls from CHB-CVS and DBDS were defined as per HERMES analysis plan using ICD-10 codes extracted from the NPR and analyzed collectively.

**DCM-Garnier^4^**

French DCM samples : CARDIGENE (n=408), EUROGENE (EHF) (n=84) and PHRC (n=204); German DCM samples : German Heart Institute Berlin cohort (n=987) and EUROGENE (EHF) study (n=214); Italian samples : EUROGENE (EHF) (n=82); French controls : PP3S study (n=1,084), German controls : KORA F4 study (n=3,264); Italian controls : EUROGENE (EHF) study (n=92)

**deCODE Heart Failure Study**

The deCODE Icelandic heart failure (HF) sample set included patients diagnosed with HF at Landspitali – The National University Hospital (LUH) in Reykjavik and the Primary Health Care Clinics of the Reykjavik capital region, following ICD-9 and ICD-10 codes for heart failure, as described in this manuscript. Information on exclusion criteria and comorbities comes from the same sources. The controls included population controls from the Icelandic genealogical database and individuals recruited through different genetic studies at deCODE genetics. Individuals of non-Icelandic origin were excluded from the study.

**DiscovEHR**

DiscovEHR is a collaboration between Geisinger and the Regeneron Genetics Center. The population is derived from patients who have previously consented to participate in the Geisinger MyCode Community Health Initiative. MyCode is an IRB-approved research study, and all participants have provided informed consent for broad use of samples for research. Participants are broadly recruited to MyCode from both primary and specialty care clinics across the Geisinger system. Exome sequencing and genome-wide genotyping of samples are performed by Regeneron, and genomic data are linked with Geisinger’s long-standing electronic health record, comprising both inpatient and outpatient records. At the time of this study, data for 144,204 patients were available for analysis. This work was supported by the Regeneron Genetics Center and Geisinger.

**Estonian Biobank (EstBiobank)^5^**

The Estonian Biobank (EstBiobank) is a population-based biobank with over 200,000 participants. All biobank participants have signed a broad informed consent form. All EstBiobank participants have been genotyped at the Core Genotyping Lab of the Institute of Genomics, University of Tartu. Phenotypes for the present analysis were defined according to HERMES 2.0 analysis plan, using electronic health records (EHRs) available to EstBiobank. Individuals were phenotyped according to 109 separate ICD-10 diagnosis codes (EHRs from Estonian Health Insurance Fund, epicrises and local hospitals) and/or drug prescriptions based on two separate ATC codes (data from Estonian Health Insurance Fund and self-reported by biobank participants). Information on ICD codes is obtained via regular linking with the national Health Insurance Fund and other relevant databases.

**The 100,000 Genomes Project (GeL)**

The 100,000 Genomes Project (GeL) is a national UK programme that recruited probands with rare diseases and cancer from clinical centres, together with family members, and performed germline and somatic (for a subset of participants with cancer) whole genome sequencing^6,7^. DCM cases were identified from HPO terms at time of study recruitment, and ICD codes from preceding and subsequent clinical episodes.

**The Genetics of Diabetes Audit and Research Tayside Scotland (GoDARTS)^8^**

GoDARTS is a cohort study of 18,306 participants, 10,149 with type 2 diabetes and 8,157 healthy controls at baseline recruited from 1996 to 2009 in Tayside, Scotland. Genetic data are available for 8,564 T2D cases and 4,586 controls. Overall, 53.33% of the cohort are male. The majority of the cohort are Caucasian (99.70%) and the median age at recruitment was 64 years. Patients consented to electronic health record linkage to allow follow-up on mortality, hospitalisations and investigations including echocardiography.

**Myocardial Applied Genomics Network (MAGNet)**

MAGNet is a collaborative group of investigators based at Perelman School of Medicine, University of Pennsylvania who use genomic approaches to understand human myocardial disease (http://www.med.upenn.edu/magnet/, NHLBI grant number 1R01HL105993-01A1). The present analysis includes a case-control GWAS comparing a subset of patients with prevalent heart failure referred for evaluation and treatment at a heart failure specialty centre at a local institution. Prevalent heart failure was identified by a heart failure cardiologist based on clinical evaluation and cardiac imaging.

**The Mass General Brigham Biobank (MGB)**

The Mass General Brigham (formerly “Partners Healthcare”) Biobank is a large research data and sample repository comprising more than 100,000 participants that is embedded within the framework of Mass General Brigham Personalized Medicine. Participants are prospectively enrolled in the context of outpatient visits, inpatient stays, and emergency department encounters. The Biobank contains banked samples (plasma, serum, DNA and buffy coats), genomic data, and other health information, including data from the electronic health record (EHR) at hospitals affiliated with the Mass General Brigham Healthcare system – primarily the Massachusetts General Hospital and the Brigham and Women’s Hospital in Boston, MA. Array- based genotyping was performed using either the Illumina Multi-Ethnic Genotyping Array, Expanded Multi-Ethnic Genotyping Array, or the Multi-Ethnic Global BeadChip Array (Illumina, Inc., San Diego, CA). The present study included the first 25,784 genotyped participants of European genetic ancestry from the Mass General Brigham Biobank with relevant clinical data available.

**Prospective Investigation of the Vasculature in Uppsala Seniors (PIVUS)^9^**

The PIVUS study, initiated in 2002, is a community-based prospective cohort comprising
1016 randomly selected men and women aged 70 years in Uppsala county. Subjects were
re-investigated at age 75 and age 80 years. The PIVUS-study is unique in its detailed
characterization of vascular and cardiac function and morphology. The cohort is also well
characterized with regards to genomics, epigenomics, proteomics and metabolomics. Ten year follow-up data on cardiovascular events and mortality is available.

**The Royal Brompton and Harefield Hospital, UK (RBH)^10^**

The Royal Brompton & Harefield Hospitals, UK are tertiary national cardiomyopathy referral centers. Participants with dilated cardiomyopathy were diagnosed using national and international clinical guidelines, with advanced cardiac imaging in all cases, and were recruited into the Royal Brompton & Harefield Hospitals Cardiovascular Research Biobank^11^ (RBH).

**University College of London, UK (UCL)**

The UCL Dilated Cardiomyopathy Cohort were recruited as part of the Inherited Cardiac Disease: A clinical and genetic investigation study, a research registry of patients referred to specialist cardiomyopathy clinics at the UCLH Heart Hospital and Saint Bartholomew’s Hospital. Patients were included who had a clinical diagnosis of DCM (defined as LVEF < 50% in the absence of abnormal loading conditions or coronary artery disease sufficient to cause the phenotype). Sample dates ranged from October 2010 to September 2020.

**UK Biobank (UKB)^12^**

The UK Biobank is a population-based prospective cohort with extensive genetic and phenotypic data collected on approximately 500,000 individuals aged 40–69 years recruited from across the UK between 2006 and 2010. Collected information include socio-demographics, lifestyle, and health-related factors, physical measures, biological samples (blood, urine, and saliva) for genomics and biochemical markers assessments, linked electronic health records, disease registers, and death register, with a planned repeat assessments and multi-modal imaging. The UK Biobank genetic data contains genotypes for 488,377 participants assayed using two very similar genotyping arrays with extensive phasing and genotype imputation. To define phenotypes used in the GWAS analysis, the present study used the diagnosis and procedure codes in linked primary care record (covering 228,948 participants with latest clinical event recorded on 18 August 2019) and hospital admission record (covering 435,632 participants with latest diagnosis or procedure code recorded on 30 June 2020).

**Uppsala Longitudinal Study of Adult Men (ULSAM)^13^**

The ULSAM study is a longitudinal community based cohort study of 50 year old men that started in 1970 (n=2322). All 50 year old men in Uppsala were invited to participate. Subjects were reinvestigated at the ages of 60, 70, 77, 82 and 88 years. GWAS data are available in approximately half of the study sample. Untargeted metabolomics and proteomics data are available both in serum and urine in subsamples of the cohort. A large number of cardiovascular events and mortality data are available with more than 20 years of follow-up.

### **Study level GWAS methods and QC**

The HERMES Consortium recommended individual studies perform GWAS using the following procedure, but did not mandate it. All individual genotyping, quality control (QC), imputation, and analysis methods are reported in **Supplementary Table 2**. Ancestries were defined using self-reported followed by validation using genetically determined ancestry. Pre-imputation quality control (QC) recommendations included per marker (genotyping rate >98%, clear Hardy Weinberg equilibrium deviations and P-values in controls >1x10^-6^, duplicate concordance, Mendelian inconsistencies, minor allele frequency [MAF] concordance with reference populations, MAF>0.5% or >1%) and per sample (genotyping rate >95%, heterozygosity within 3 standard deviations, exclusion of genetic ancestry outliers on principal component [PC] plots) checks. Related individuals (pihat >0.1) were excluded unless specialized models that account for relatedness (e.g. mixed linear models) were used. Post-QC genotypes were imputed using the Michigan Imputation Server with HRC r1.1 2016 reference. GWAS was performed per study per phenotype, using logistic regression assuming additive genetic effects, adjusted for age, sex, genetic principal components (PC), and study-specific covariates. Additional study-level QC was performed for GWAS summary statistics: variants with >2 alleles, regression coefficients >10, coefficient standard errors >10, minor allele frequency (MAF) <1%, imputation (INFO) score <0.6, or effective sample size <50 were excluded. Of remaining variants, allele frequency differences >0.2 compared against European ancestry reference panel (whole-genome sequencing data from HRC and 1000G project phase 3 version 5) were further excluded. Genomic inflation adjustment were applied for summary statistics with genomic control coefficient >1.1.

### **Published dilated and non-ischaemic cardiomyopathy GWAS**

| **Study first author** | **Year** | **Phenotype** | **Cases** | **Controls** | **Reported lead variants** | | | | |
| --- | --- | --- | --- | --- | --- | --- | --- | --- | --- |
|  |  |  |  |  | **Variant** | **Position** | **EA**  **(Freq)** | **OR** | **Annotated gene** |
| Garnier^4^ | 2021 | DCM | 2651 | 4329 | rs62232870  rs4684185  rs148248535  rs7284877 | 3:14257709  3:14272914  16:2183449  22:24155111 | A (0.23)  C (0.7)  T (0.82)  C (0.81) | 1.36  1.28  1.35  1.32 | *LSM3*  *LSM3*  *PDKD1*  *SMARCB1* |
| Tadros^14^ | 2021 | DCM MTAG | 5521 | 397323 | rs10927875  rs2291569  rs2234962  rs2042995  rs6807275  rs4713999  rs13265989  rs12541595  rs2182400  rs1051168  rs9892651  rs2303510  rs2186370 | 1:16299312  7:128488734  10:1214296332:179558366  3:14274451  6:36633069  8:11779640  8:125857359  10:29707551  15:85200520  17:64303793  18:34324091  22:24171305 | C (0.67)  G (0.92)  T (0.78)  T (0.78)  G (0.66)  G (0.67)  G (0.29)  G (0.69)  G (0.24)  T (0.26)  C (0.42)  G (0.68)  G(0.81) | 1.14  1.16  1.21  1.11  1.08  1.11  1.09  1.10  1.10  1.09  1.08  1.08  1.12 | *ZBTB17*  *FLNC*  *BAG3*  *TTN*  *LSM3*  *CDKN1A*  *CTSB*  *MTSS1*  *SVIL*  *NMB*  *PRKCA*  *FHOD3*  *SMARCB* |
| Aragam^15^ | 2018 | NICM | 2038 | 388326 | rs2234962  rs2634071  rs6843082  rs12138073  rs34471231  rs10927875 | 10:1214296334:111669220  4:111718067  1:16354985  1:16356522  1:16299312 | T (0.78)  T (0.16)  G (0.19)  T (0.10 G (0.10)  C (0.68) | 1.3  1.25  1.24  1.29  1.29  1.15 | *BAG3*  *PITX2*  *PITX2*  *CLCNKA*  *CLCNKA*  *ZBTB17* |
| Esslinger^16^ | 2017 | DCM | 2344* | 2410* | rs10927875  rs2234962 | 1:1617899  10:121419623 | C (0.68)  T (0.78) | 1.47  1.56 | *ZBTB17*  *BAG3* |
| Meder^17^ | 2014 | DCM | 4100** | 7600** | rs9262636 | 6:31025848 | G (0.22) | 1.48 | *HCG22* |
| Villard^18^ | 2011 | DCM | 2344** | 2410** | rs10927875  rs2234962 | 1:16171899  10:12141962 | C (0.34)  C (0.21) | 0.71  0.64 | *ZBTB17*  *BAG3* |

*1 replication cohort

**2 replication cohorts

EA = effect allele; OR = odds ratio; DCM = dilated cardiomyopathy; NICM = non-ischemic cardiomyopathy

**Curation of pathogenic and likely pathogenic rare variants in DCM-causing genes in the UK Biobank**

Variants 100 bp up- and down-stream the region of 19 DCM-associated genes with definitive, strong, or moderate evidence for disease, were extracted from the whole exome sequencing data of 454,756 UKB participants^19^. MANE, protein-altering variants that had a MAF <0.1% in gnomAD and UKBB were identified. Intron variants that were pathogenic in ClinVar were manually curated for functional evidence of splicing. Splice region variants and other splice variants (excluding essential splice and splice donor 5th base), were included if they were predicted to cause splicing through SpliceAI (threshold >0.8)^20^. Only *TTN* truncating variants in PSI>90% exons or variants with a splice prediction or curation of pathogenic (with assessment) were included.

The variant list was then shortened to only include the 12 definitive or strong evidence DCM genes (*BAG3*, *DES*, *DSP*, *FLNC*, *LMNA*, *MYH7*, *PLN*, *RBM20*, *SCN5A*, *TNNC1*, *TNNT2*, *TTN*). LOFTEE was used to identify low confidence LoF variants and identify PAVs that are mislabelled (e.g. NAGNAG sites)^21^. All predicted LoF variants were annotated for prediction of NMD escaping; using the NMD plugin, a threshold of 55bp into the penultimate exon of the genes, and additional curation of frameshift variants upstream to the 55bp threshold but where the PTC is predicted to be within the threshold using HGVSp. Additionally, "missense_variant, splice_region_variant"s were only flagged as NMD escaping if they also had a prediction of splicing by SpliceAI. The variants were then filtered for disease-causing mechanisms; all protein-altering variants were kept for *BAG3*, *LMNA*, *PLN*, *SCN5A*, *RBM20*, and *DSP*; variants influencing gene product structure (i.e. indels, missense, start and stop lost) or gene product level (i.e. frameshift, splice, stop gained) (but NMD escaping) were kept for *MYH7*, *DES*, *TNNC1*, and *TNNT2*; and variants predicted to influence gene product level were kept for *TTN* and *FLNC*.

The variant list was then shortened to only include variants that met a filtering allele frequency (FAF) (gnomAD population max FAF <0.00004) and as a MAF in UKB. Additionally, compound heterozygous carriers of common TTN truncating variants (same two TTN variants identified in >10 individuals) were removed from the analysis as these were likely rescued. Finally, only variants that would be called pathogenic or likely pathogenic if identified in a patient with DCM were included; using Cardioclassifier and ClinVar, the variants were manually curated if they had any evidence of pathogenicity.

**Individual study acknowledgements**

***Bio-SHiFT-TRIUMPH and Doetinchem Cohort Study***

The Bio-SHiFT study was supported by the Jaap Schouten Foundation and the Noordwest Academie. TRIUMPH was supported by the framework of the Center for Translational Molecular Medicine, project TRIUMPH (grant 01C-103).

We thank the Dutch Ministry of Health, Welfare and Sport and the National Institute for Public Health and the Environment (RIVM), Bilthoven, the Netherlands, for their contribution and ongoing support to the Doetinchem Cohort Study

***BioVU***

The dataset(s) used for the analyses described were obtained from Vanderbilt University Medical Center’s BioVU which is supported by numerous sources: institutional funding, private agencies, and federal grants. These include the NIH funded Shared Instrumentation Grant S10RR025141; and CTSA grants UL1TR002243, UL1TR000445, and UL1RR024975.

***CHB***

We would like to thank the patients in CHB-CVDC and the participants in DBDS. We thank deCODE genetics/Amgen and Department of Clinical Immunology, Copenhagen University Hospital for financial support of this study. Furthermore, we thank the staff working in the biobank facilities and in the hospitals whose work make the existence of this cohort sustainable. This work was supported by the Novo Nordisk Foundation (NNF) (grant numbers: NNF17OC0027594, NNF14CC0001 and NNF18SA0034956), Innovation Fund Denmark (grant number: 5153-00002B) and Nordforsk, Precision Medicine Heart (grant number: 90580 PM Heart).

***deCODE***

We at deCODE thank the women and men of Iceland that have participated in our studies and our colleagues that contributed to data collection and processing.

***DiscovEHR***

We acknowledge and thank all participants in Geisinger’s MyCode Community Health Initiative for their support and permission to use their health and genomic information in the DiscovEHR collaboration. This work was supported by the Regeneron Genetics Center and Geisinger.

***EstBiobank***

The establishment and maintenance of the Estonian Biobank has been supported by Targeted Financing from the Estonian Ministry of Science and Education [SF0180142s08], the Ministry of Social Affairs and the Ministry of Economic Affairs and Communications; the biobank and the Genome Center have been supported by the Development Fund of the University of Tartu (grant SP1GVARENG); the European Regional Development Fund has supported the Centre of Excellence in Genomics (EXCEGEN; grant 3.2.0304.11-0312) and through FP7 grants [278913, 306031, 313010, 245536, 269213, 212111, 201413].

***Garnier-DCM***

This work was supported by grants from the GENMED Laboratory of Excellence on Medical Genomics [ANR-10-LABX-0013]; DETECTIN-HF project (ERA-CVD framework); Assistance Publique-Hôpitaux de Paris [PHRC programme hospitalier de recherche Clinique, AOM04141]; Délégation à la recherche clinique AP-HP [EMUL and PHRCn_AOM95082]; the ‘Fondation LEDUCQ’ [Eurogene Heart Failure network; 11CVD 01]; the PROMEX charitable foundation and the Société Française de Cardiologie/Fédération Française de Cardiologie. The SFBTR19 registry was supported by the Deutsche Forschungsgemeinschaft (DFG). The KORA study was initiated and financed by the Helmholtz Zentrum München – German Research Center for Environmental Health, funded by the German Federal Ministry of Education and Research (BMBF) and by the State of Bavaria. KORA research was supported at the Munich Center of Health Sciences (MC-Health), Ludwig-Maximilians-Universität, as part of LMUinnovativ.

***GeL***

This research was made possible through access to the data and findings generated by the 100,000 Genomes Project under Project Number 667. The 100,000 Genomes Project is managed by Genomics England Limited (a wholly owned company of the Department of Health and Social Care), and funded by the National Institute for Health Research and NHS England. The Wellcome Trust, Cancer Research UK and the Medical Research Council have also funded research infrastructure. The 100,000 Genomes Project uses data provided by patients and collected by the National Health Service as part of their care and support. Genotyping was supported by the Institute of Psychiatry Psychology and Neuroscience (IoPPN) Genomics & Biomarker Core Facility within King's College London, who gratefully acknowledge capital equipment funding from the Maudsley Charity [980] and Guy’s and St Thomas’s Charity [STR130505].

***MAGNet***

We thank the Broad Institute for generating high-quality sequence data with Patrick Ellinor as the PI. The datasets used in this manuscript were obtained from dbGaP at http://www.ncbi.nlm.nih.gov/gap through dbGaP accession number phs001539.

***RBH***

This work was supported by funding from the British Heart Foundation [RE/18/4/34215, FS/IPBSRF/22/27059, FS/15/81/31817, FS/ICRF/21/26019, RG/19/6/34387, BC/F/21/220106]; the Medical Research Council [MC_UP_1605/13]; Wellcome Trust [107469/Z/15/Z]; the National Institute for Health Research (NIHR) Imperial College Biomedical Research Centre; NIHR Royal Brompton Cardiovascular Biomedical Research Unit; Sir Jules Thorn Charitable Trust [21JTA]; National Heart Lung Institute Foundation; Royston Centre for Cardiomyopathy Research; Rosetrees Trust. This research has been conducted in part using the UK Biobank Resource under Application Numbers 9922, 15422, 18545, 40616 and 47602.

**University College of London, UK (UCL)**

The authors are grateful to the patients and their families whose contributions have made this work possible. This study was supported by the National Institute for Health Research University College London Hospitals Biomedical Research Centre.

***UK Biobank***

UK Biobank was established by the Wellcome Trust medical charity, Medical Research Council, Department of Health, Scottish Government and the Northwest Regional Development Agency. It has also had funding from the Welsh Government, British Heart Foundation, Cancer Research UK and Diabetes UK. UK Biobank is supported by the National Health Service (NHS). UK Biobank has received additional funding for: genotyping of all 500,000 participants (from the Department of Health, Medical Research Council, British Heart Foundation); for the measurement of biochemistry markers in all 500,000 participants (from the Wellcome Trust, Medical Research Council, British Heart Foundation, Diabetes UK); for imaging 100,000 participants (Wellcome Trust, Medical Research Council, British Heart Foundation) and re-imaging 10,000 participants (Dementia Platform UK), for the whole genome sequencing of the full cohort (Medical Research Council, and for establishing a research analysis platform (Wellcome), and for repeat imaging up to 60,000 participants from the imaging programme (Medical Research Council, CZI and Calico).

***ULSAM***

This study was supported by grants from Primary Health Care in Uppsala County, Royal Scientific Society Foundation (Kungliga vetenskapssamhällets fond), Swedish Heart Lung Foundation (Hjärt-Lungfonden) and Thuréus Foundation.

**HERMES Consortium Contributors**

Erik Abner, Lance Adams, Peter Almgren, Charlotte Andersson, Krishna G. Aragam, Johan Ärnlöv, Folkert W. Asselbergs, Geraldine Asselin, Joshua D. Backman, John Baksi, Paul J.R. Barton, Traci M. Bartz, Traci Bartz, Kiran J. Biddinger, Mary L. Biggs, Heather L. Bloom, Eric Boersma, Isabelle Bond, Maxime Bos, Jeffrey Brandimarto, Michael R. Brown, Hans-Peter Brunner-La Rocca, Rachel Buchan, Leonard Buckbinder, Douglas Cannie, Thomas P. Cappola, David J. Carey, Mark D. Chaffin, Philippe Charron, Daniel I. Chasman, Xing Chen, Sandesh Chopade, William Chutkow, John G.F. Cleland, James P. Cook, Stuart A. Cook, Tomasz Czuba, Trégouët David-Alexandre, Simon Dedenus, Abbas Dehghan, Graciela E. Delgado, Spiros Denaxas, Alexander S. Doney, Marie-Pierre Dubé, J. Gustav Smith, Samuel C. Dudley, Michael E. Dunn, Marcus Dörr, Patrick T. Ellinor, Perry Elliott, Gunnar Engström, Villard Eric, Tõnu Esko, Eric H. Farber-Eger, Eric Farber-Eger, Stephan B. Felix, Chris Finan, Sarah Finer, Ian Ford, René Fouodjio, Catherine Francis, Sophie Garnier, Mohsen Ghanbari, Sahar Ghasemi, Jonas Ghouse, Vilmantas Giedraitis, Franco Giulianini, John S. Gottdiener, Jasmine Gratton, Stefan Gross, Hongsheng Gui, Karl Guo, Rebecca Gutmann, Daníel F. Guðbjartsson, Christopher M. Haggerty, Brian Halliday, Åsa K. Hedman, Anna Helgadottir, Harry Hemingway, Albert Henry, Hans Hillege, Aroon D. Hingorani, Hilma Holm, Craig L. Hyde, Sara Hägg, Hanane Issa, Jaison Jacob, Deleuze Jean-François, J. Wouter Jukema, Frederick Kamanu, Isabella Kardys, Ravi Karra, Maryam Kavousi, Kay-Tee Khaw, Jorge R. Kizer, Jorge Kizer, Marcus E. Kleber, Andrea Koekemoer, Bill Kraus, Karoline Kuchenbaecker, Lars Køber, Yi-Pin Lai, David Lanfear, Chim C. Lang, Claudia Langenberg, Michael Lee, Honghuang Lin, Lars Lind, Cecilia M. Lindgren, Peter P. Liu, Barry London, Brandon D. Lowery, Brandon Lowery, Jian'an Luan, Steven A. Lubitz, R. Thomas Lumbers, Patrik Magnusson, Anubha Mahajan, Anders Malarstig, Douglass Mann, Kenneth B. Margulies, Nicholas A. Marston, Nicholas Marston, Hillary Martin, Kathryn McGurk, John J.V. McMurray, Olle Melander, Giorgio Melloni, Andres Metspalu, David Miller, Xiaodong Mo, Ify R. Mordi, Michael P. Morley, Andrew P. Morris, Andrew D. Morris, Alanna C. Morrison, Lori Morton, Winfried März, Michael W. Nagle, Christopher P. Nelson, Alexander Niessner, Teemu Niiranen, Raymond Noordam, Michela Noseda, Christoph Nowak, Michelle L. O'Donoghue, Declan P O'Regan, Anjali T. Owens, Colin N. A. Palmer, Antonis Pantazis, Helen M. Parry, Guillaume Paré, Markus Perola, Louis-Philippe Lemieux Perreault, Charron Philippe, Marie Pigeyre, Eliana Portilla-Fernandez, Sanjay K. Prasad, Bruce M. Psaty, Kenneth M. Rice, Paul M. Ridker, Simon P. R. Romaine, Carolina Roselli, Jerome I. Rotter, Christian T. Ruff, Mark S. Sabatine, Mark Sabatine, Neneh Sallah, Perttu Salo, Veikko Salomaa, Nilesh J. Samani, Naveed Sattar, A. Floriaan Schmidt, Jessica van Setten, Sonia Shah, Svati Shah, Alaa A. Shalaby, Akshay Shekhar, Diane T. Smelser, Nicholas L. Smith, J. Gustav Smith, Garnier Sophie, Doug Speed, Sundararajan Srinivasan, Kari Stefansson, Steen Stender, David J. Stott, Garðar Sveinbjörnsson, Per Svensson, Daniel I. Swerdlow, Petros Syrris, Mari-Liis Tammesoo, Jean-Claude Tardif, Upasana Tayal, Maris Teder-Laving, Alexander Teumer, Pantazis I Theotokis, Guðmundur Thorgeirsson, Unnur Thorsteinsdottir, Christian Torp-Pedersen, Vinicius Tragante, Stella Trompet, Andre G. Uitterlinden, Ramachandran S. Vasan, Felix Vaura, Abirami Veluchamy, Monique Verschuuren, Niek Verweij, Adriaan A. Voors, Marion van Vugt, Uwe Völker, Lars Wallentin, Yunzhang Wang, James S. Ware, James Ware, Nicholas J. Wareham, Dawn Waterworth, Peter E. Weeke, Peter Weeke, Raul Weiss, Quinn Wells, Harvey White, Kerri L. Wiggins, Jemma B. Wilk, L. Keoki Williams, Xiao Xu, Yifan Yang, Laura M. Yerges-Armstrong, Bing Yu, Faiez Zannad, Jing Hua Zhao, Sean L. Zheng, Simon de Denus, Antonio de Marvao, David van Heel, Jessica van Setten, Marion van Vugt, Pim van der Harst

**Supplementary Method References**
