## Supplementary Figure for "Genome-wide association analysis reveals insights into the molecular etiology underlying dilated cardiomyopathy"

### Supplementary Figures

**Figure S1**: QQ plots for $GWAS_{DCM}$, $GWAS_{MTAG}$ and $GWAS_{DCM\text{-}strict}$

**Figure S2:** Manhattan plot of $GWAS_{DCM\text{-}strict}$

**Figure S3**: Forest plot of effect size across $GWAS_{DCM}$, $GWAS_{MTAG}$ and $GWAS_{DCM\text{-}strict}$

**Figure S4**: Scatter plot of absolute effect sizes for conditionally independent variants in $GWAS_{DCM}$ and $GWAS_{DCM\text{-}Strict}$, and in $GWAS_{DCM}$ and $GWAS_{MTAG}$.

**Figure S5**: Upset plot of overlapping and uniquely significant loci in $GWAS_{DCM}$ (FDR<1%), $GWAS_{DCM\text{-}Strict}$ ($P<5x10^{-8}$) and $GWAS_{MTAG}$ ($P<5x10^{-8}$)

**Figure S6**: Functionally-informed fine-mapped variants at genomic loci

**Figure S7**: Summary of candidate gene prioritization results

**Figure S8**: Integration of genomics and single-nuclei transcriptomics identifies genes and biological mechanisms important in DCM

**Figure S9**: Intercellular communication inferred from single-nuclei transcriptomics

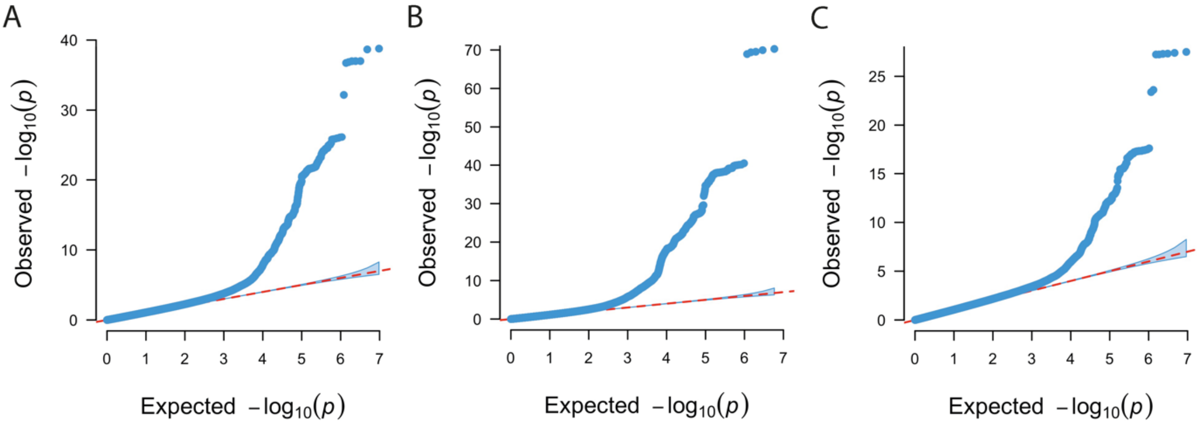


**Figure S1**: Quantile-quantile plots for (A) $GWAS_{DCM}$, (B) $GWAS_{MTAG}$ and (C) $GWAS_{DCM\text{-}strict}$.

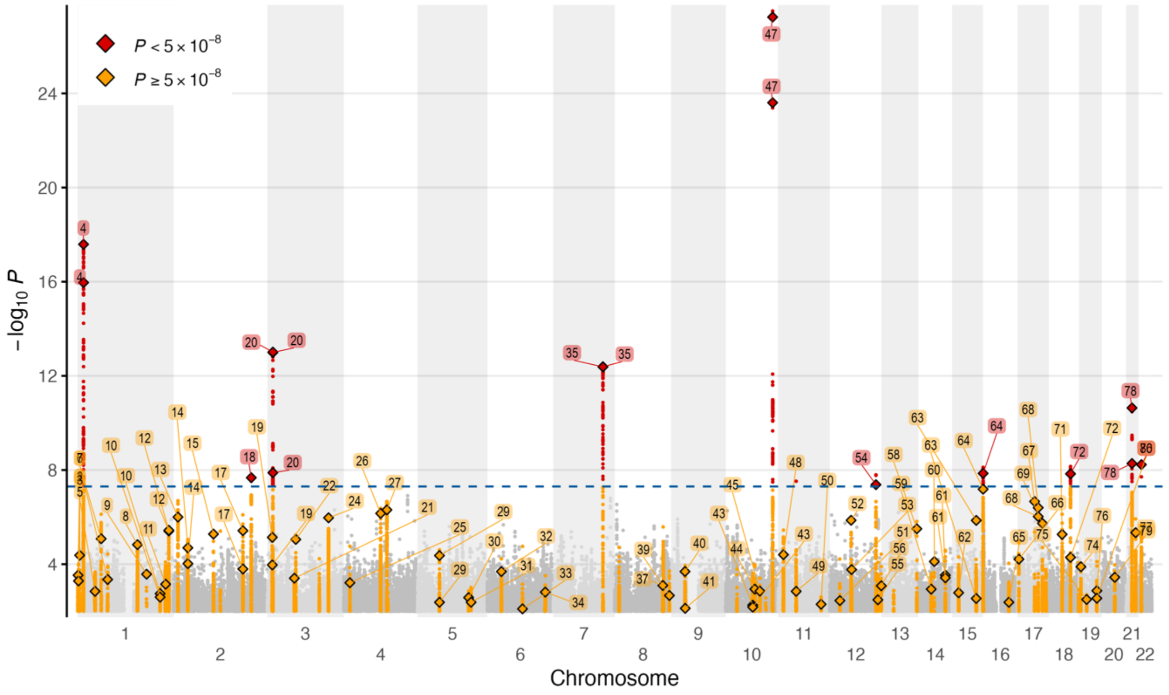


**Figure S2**: Manhattan plot of GWAS of 6,001 strictly defined DCM cases and 449,384 controls ($GWAS_{DCM\text{-}Strict}$). The 80 loci identified from $GWAS_{DCM}$ and $GWAS_{MTAG}$ (Figure 2) are labelled. DCM diagnosis required cardiac imaging, clinical expertise and/or robustly-defined ICD codes. In total there were 10 loci reaching genome-wide significance (dashed blue line – $P<5x10^{-8}$), all of which were significant in the primary GWAS.

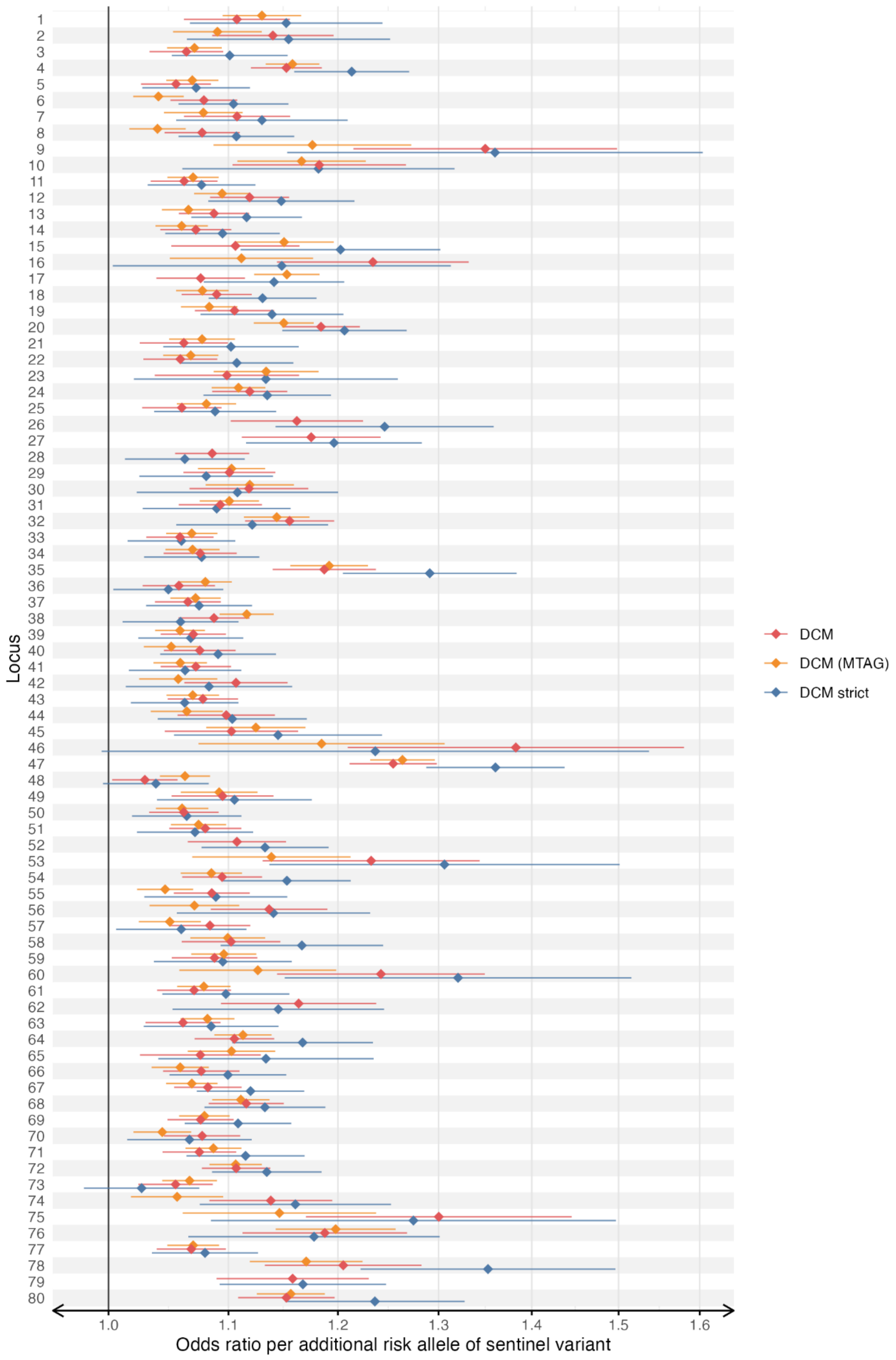


**Figure S3**: Forest plot of effect size across $GWAS_{DCM}$, $GWAS_{MTAG}$ and $GWAS_{DCM\text{-}Strict}$ for all 80 genomic risk loci identified in $GWAS_{DCM}$ and MTAG.

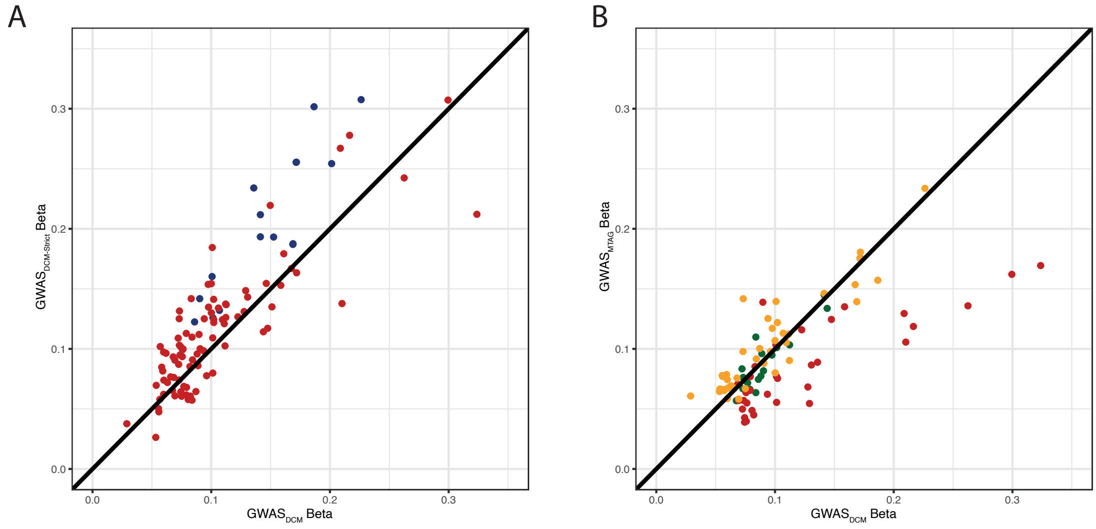


**Figure S4**: Scatter plot comparing absolute effect sizes for conditionally independent variants in (A) GWAS$_{DCM}$ and GWAS$_{DCM\text{-}Strict}$; and in (B) GWAS$_{DCM}$ and GWAS$_{MTAG}$. Variants tended to have a greater effect in GWAS$_{DCM\text{-}Strict}$ than in GWAS$_{DCM}$, particularly for variants that were genome-wide significant in GWAS$_{DCM\text{-}Strict}$ (**blue**) compared with those that were only FDR significant in GWAS$_{DCM}$ (**red**). When comparing GWAS$_{DCM}$ and GWAS$_{MTAG}$, variants that were FDR significant in GWAS$_{DCM}$ and genome-wide significant in GWAS$_{MTAG}$ (**dark green**), and that were genome-wide significant only in GWAS$_{MTAG}$ (**yellow**), had similar effect sizes, while variants that were only FDR significant in GWAS$_{DCM}$ (**red**) tended to have larger effects in GWAS$_{DCM}$ than in GWAS$_{MTAG}$.

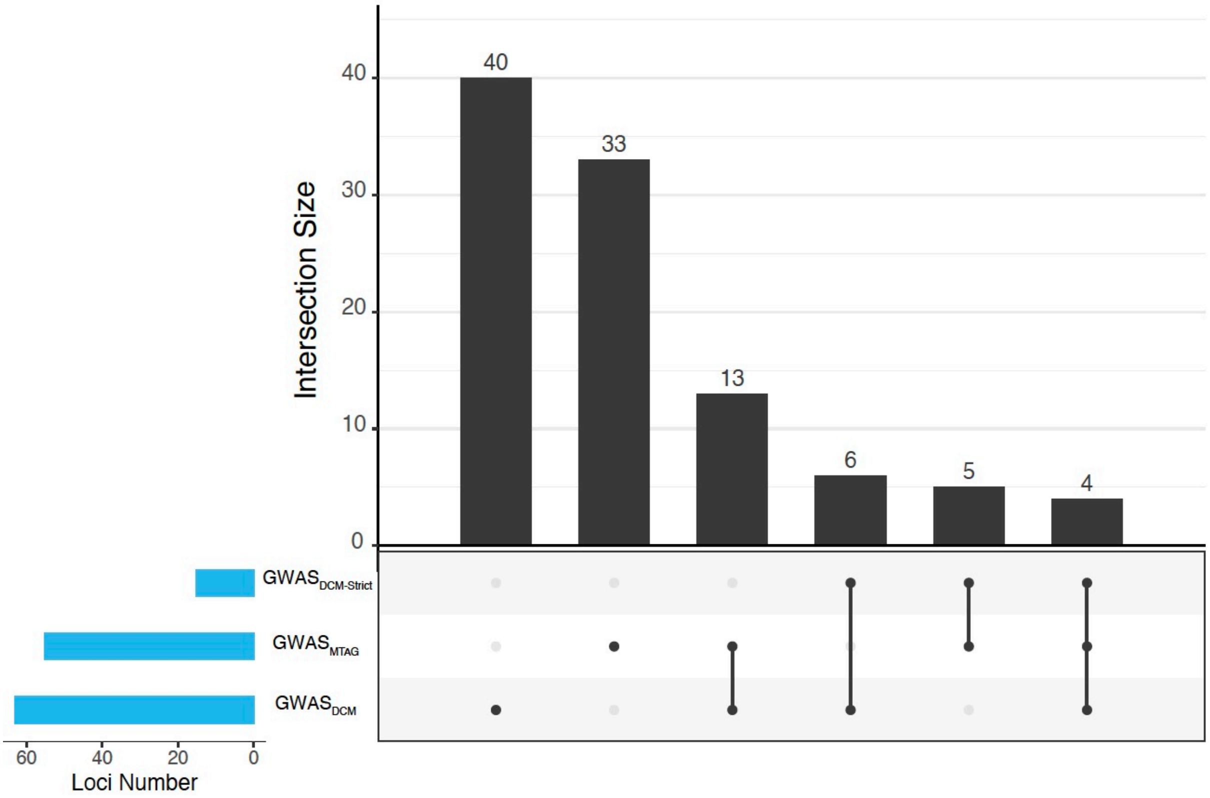


**Figure S5**: Upset plot of overlapping and uniquely significant loci in $GWAS_{DCM}$ (FDR<1%), $GWAS_{DCM\text{-}Strict}$ ($P<5x10^{-8}$) and $GWAS_{MTAG}$ ($P<5x10^{-8}$).

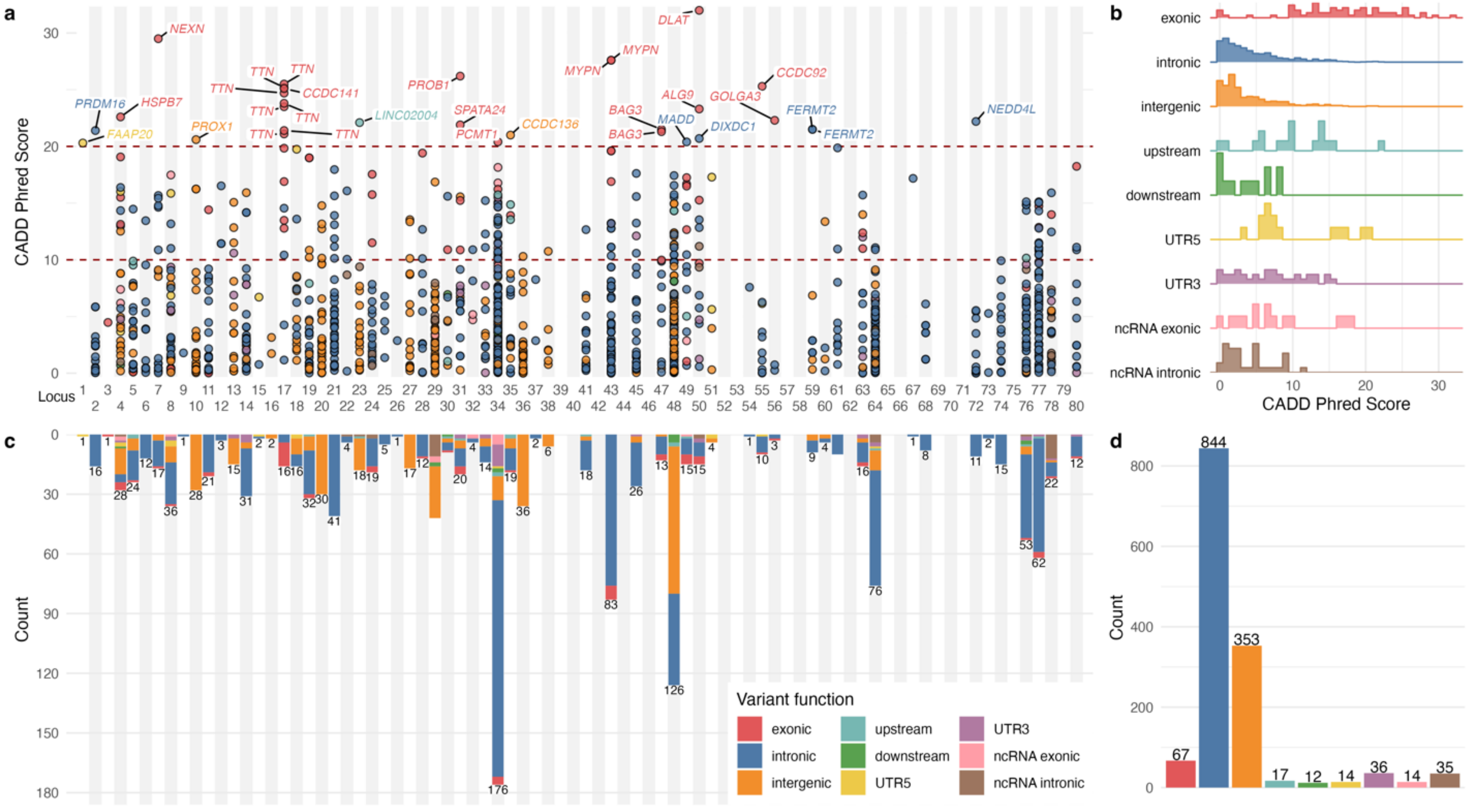


**Figure S6**: Functionally-informed fine-mapped variants at genomic loci. (A) Fine-mapped variants at genomic risk loci with variants with high CADD Phred scores (>20) annotated to the nearest gene. (B) Total number and function of fine-mapped variants at each locus. (C) Distribution of CADD Phred scores for fine-mapped variants across all genomic risk loci, stratified by variant function. (D) Number of fine-mapped variants stratified by function.

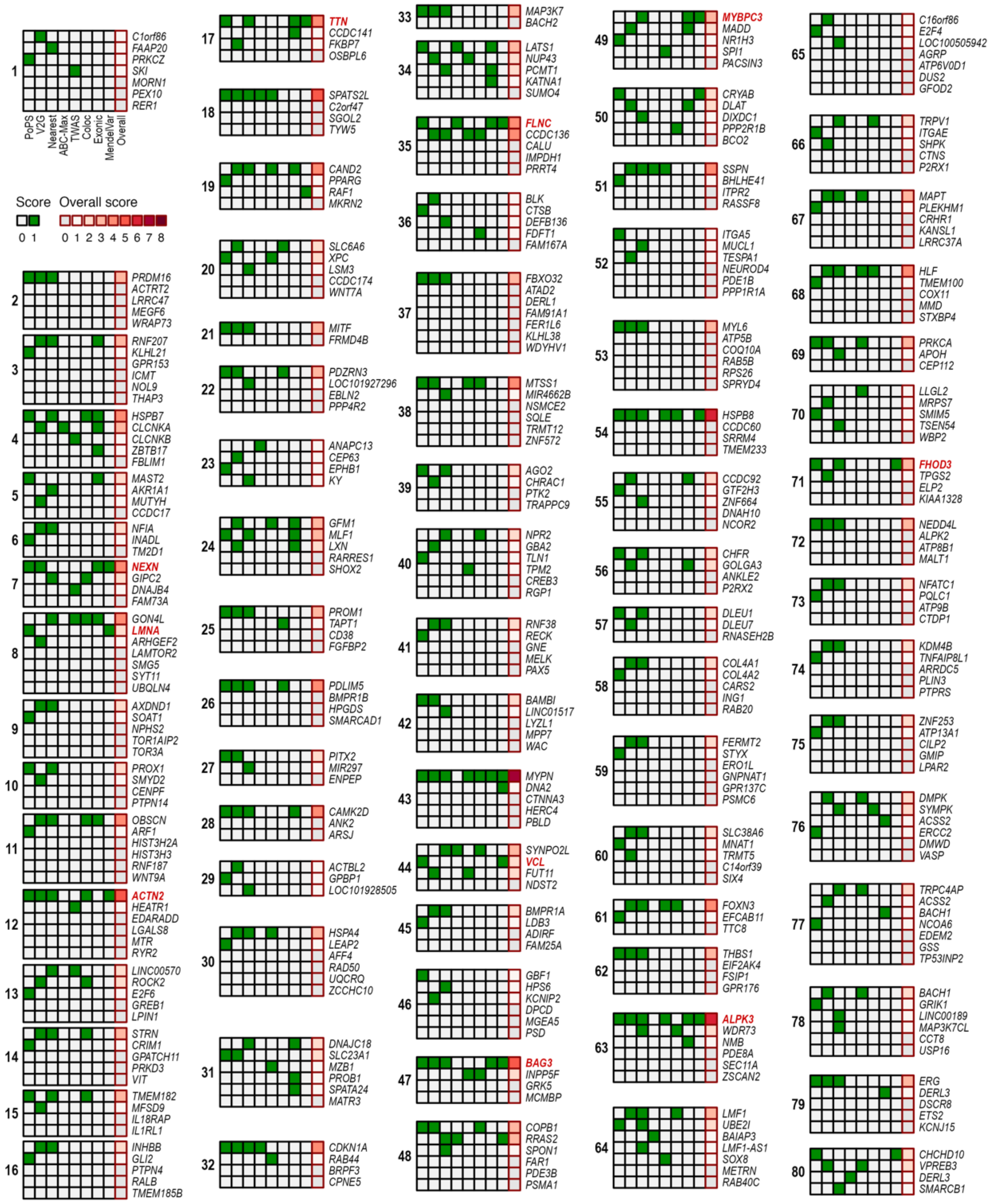


**Figure S7**: Summary of effector gene prioritization results. A two-step approach was used to identify candidate genes and prioritize potential effector genes at each loci. First, the nearest gene along with the top 3 genes scored using each of PoPs and V2G were highlighted as candidate genes for further evaluation. Second, of these genes, 5 additional features and methods were used to score the overall level of evidence supporting each putative gene by giving one point for any gene that was identified as best from each feature (maximum score of 8), and the highest scoring gene(s) at each locus being identified as the candidate gene(s). The 8 features were: PoPs, V2G, nearest, activity-by-contact (ABC)-model, transcriptome-wide association study (TWAS), colocalization, exonic coding variant, and reported Mendelian cause of cardiomyopathy or muscle disorder. Highlighted in red are genes with moderate or definitive evidence of being Mendelian causes of cardiomyopathy from ClinGen curation.

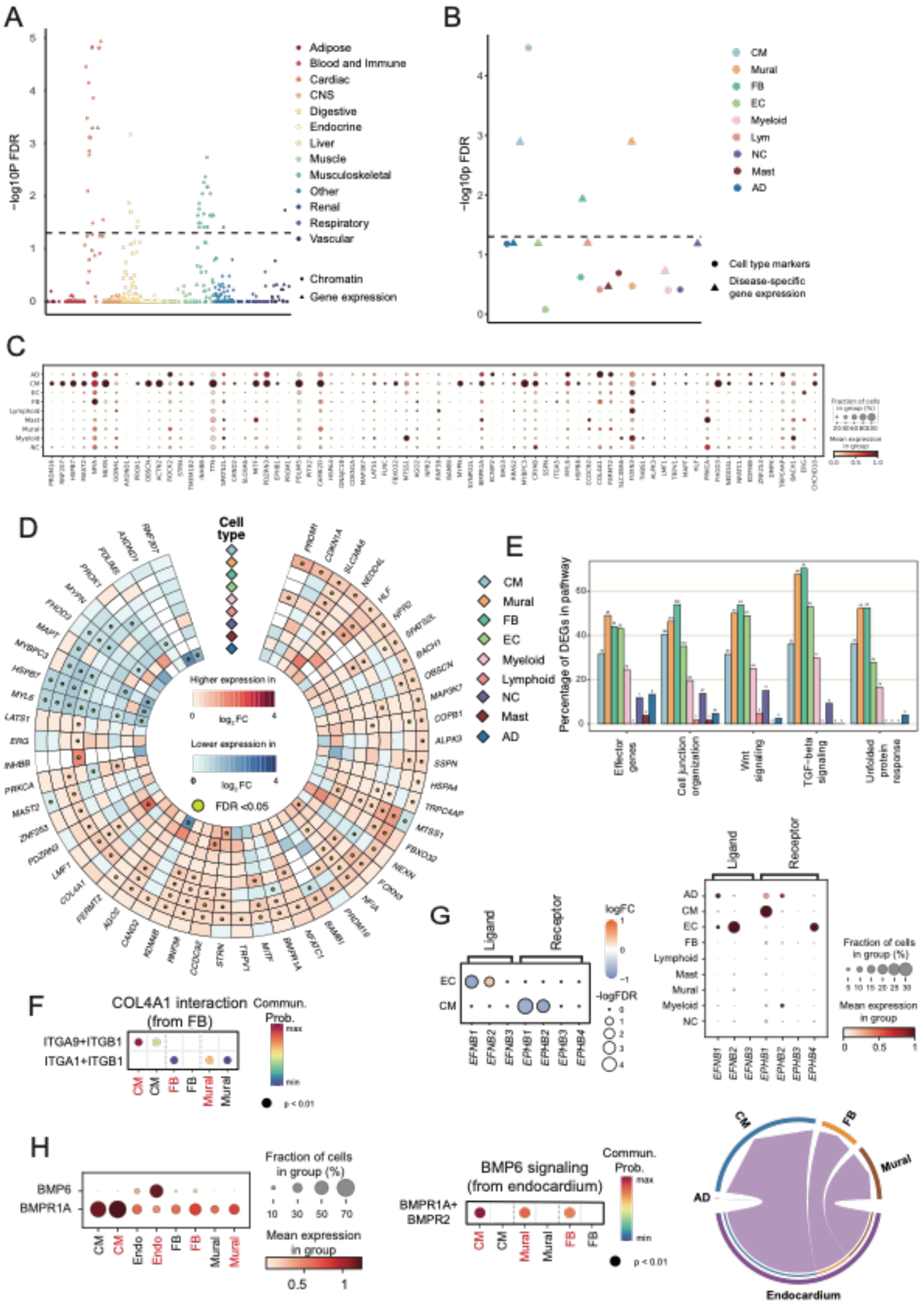


**Figure S8: Integration of genomics and single-nuclei transcriptomics identifies genes and biological mechanisms important in DCM.** Partitioned heritability at tissue level (A) and at cell type level from single-nuclei RNA-sequencing data of 52 DCM cases and 18

controls (B). (C) Cell type expression of prioritized genes in single-nuclei transcriptomics from left ventricular tissue in 18 control donors. (D) Differential expression of candidate genes across the range of cell types and states. Red and blue indicate increased and reduced gene expression in DCM compared with controls. Yellow dot indicates significant DEGs in at least one cell state within a cell type at FDR of 0.05. (E) Proportion of genes within effector gene enriched pathways that are differentially expressed in DCM compared with controls. (F) Increased COL4A1 signaling from fibroblasts to cardiomyocytes, fibroblasts, and mural cells from DCM single-nuclei transcriptomics. Communication probability indicates the scaled strength of interaction from maximum to minimum signaling interactions between cell types. (G) Increased expression of *EFNB2* (ligand) in endothelial cells and decreased expression of *EPHB1* (receptor) in cardiomyocytes in DCM. (H) Upregulation of *BMP6* (ligand) in endocardial cells resulting in increased signaling through *BMPR1A* in cardiomyoctes, fibroblasts and mural cells. Chord plot showing that majority of endocardial (purple) BMP6-BMPR1A signaling is to cardiomyocytes (blue), followed by mural (brown) and fibroblasts (orange).

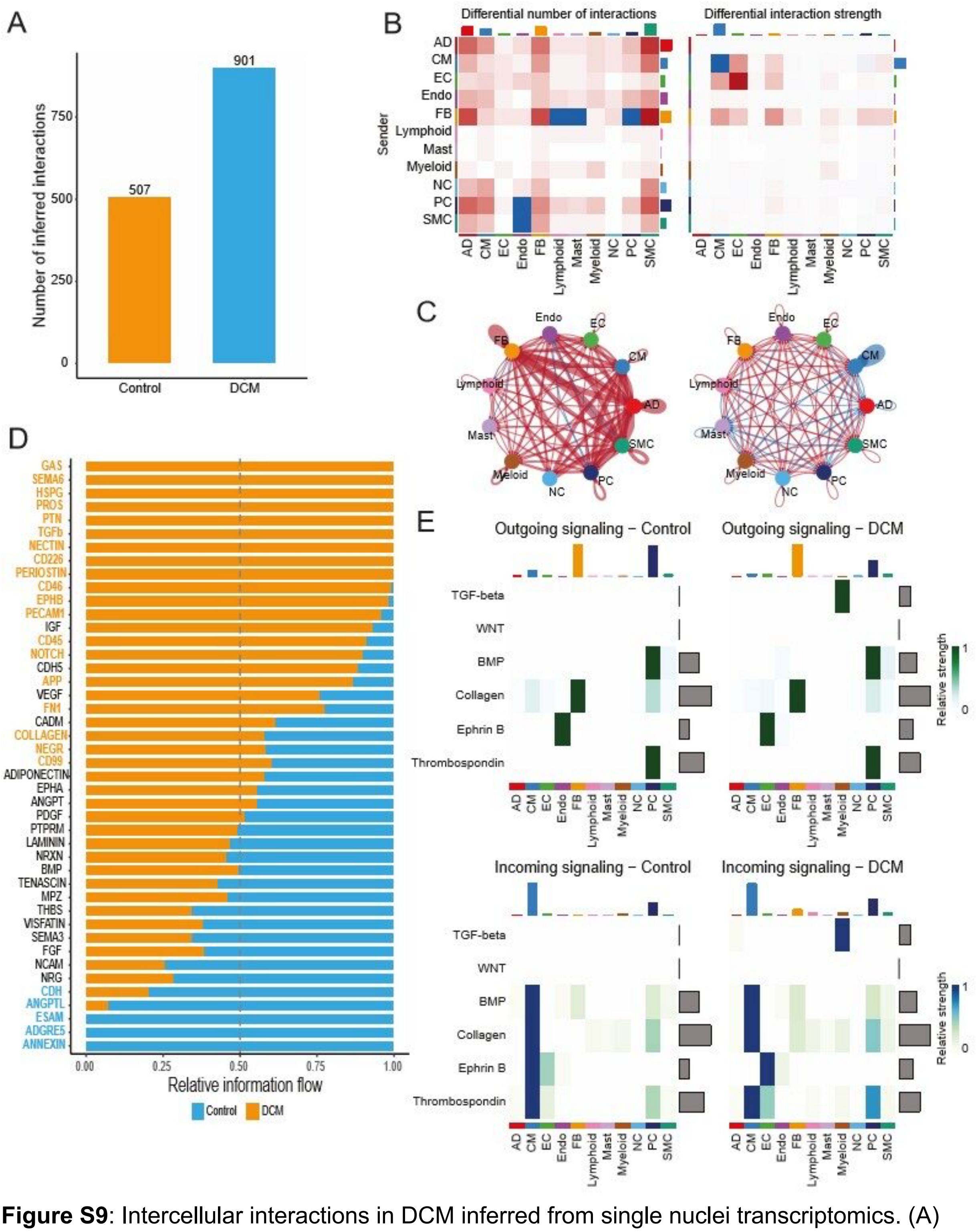


**Figure S9**: Intercellular interactions in DCM inferred from single nuclei transcriptomics. (A) Total number of interactions between cell types in DCM (blue) and control (orange). Heat map (B) and chord plot (C) showing DCM changes in intercellular communication (red – increased in DCM, blue – decreased) quantity and strength. (D) Relative information flow of curated receptor-ligand intercellular, highlighting pathways that are significantly increased in DCM (orange) or control (blue). (E) Heat map showing outgoing (green) and incoming (blue) signals for prioritized gene enriched pathways (TGF-beta and WNT pathways) and specific pathways of prioritised genes (BMP, Collagen, Ephrin B and thrombospondin). AD – adipocyte; CM – cardiomyocyte; EC – endothelial cell; Endo – endocardial cell; FB – fibroblast; NC – neuronal cell; PC – pericyte; and SMC – smooth muscle cell.
